# A first-in-human, Phase 1/2a, open-label study of SQ3370, a first-in-class doxorubicin-based click chemistry therapeutic, in patients with advanced solid tumors

**DOI:** 10.1101/2025.06.02.25328445

**Authors:** S. P. Chawla, E. Abella, S. Wieland, T.-H. Nguyen, S. Srinivasan, M. Alečković, V. Kwatra, V. Subbiah, N. Bui, V. A. Bhadri, M. C. Weiss, M. Agulnik, C. W. Ryan, A. D. Guminski, J. M. Mejía Oneto

## Abstract

**Background:** We present the first clinical proof of concept using click chemistry to selectively capture drugs at tumors. SQ3370 combines a clickable pre-targeting agent (intratumorally injected biopolymer, SQL70) and a chemically-attenuated doxorubicin (Dox) protodrug (SQP33) that is activated upon clicking with the biopolymer at the tumor to rapidly release high local concentrations of native doxorubicin.

**Methods:** This was a Phase 1/2a open-label study in patients with advanced solid tumors (NCT04106492). SQ3370 treatment comprises of an intratumoral SQL70 biopolymer injection followed by 3 or 5 consecutive daily infusions of SQP33 Dox protodrug. The primary endpoints were treatment-emergent adverse events, dose-limiting toxicities (DLTs), and to identify the recommended Phase 2 dose (RP2D). Secondary endpoints included pharmacokinetics, efficacy, and immune profiling.

**Results:** Phase 1 enrolled 39 patients. SQ3370 administered at 0.38x to 15x the standard Dox dose was well tolerated with no DLTs reported, and mild and manageable myelosuppression observed. Rapid release of Dox was observed at all the dose levels tested, with increasing exposure up to 15x the standard Dox dose; 12x was selected as the RP2D. Phase 2a enrolled 14 soft tissue sarcoma patients at 12x Dox. There was an unconfirmed objective response rate (ORR) of 14.3% (2/14 patients) and disease control rate of 71.4% (10/14; 95% CI: 41.9, 91.6). Immune profiling revealed anti-tumor immune responses, including expansion/activation of cytotoxic CD8^+^ T-cells. The study was terminated as the prespecified criteria for study continuation of ORR greater than that of standard Dox was not met.

**Conclusions:** SQ3370 is a first-in-class click chemistry-enabled, pre-targeting therapeutic, and the first reported use of *in vivo* click chemistry in humans. This approach enabled high Dox concentrations at the tumor, with minimal off-target toxicity, unlocking favorable immune responses. Objective clinical activity was observed, but ORR was comparable to standard Dox.

**Highlights:**

▪ SQ3370 is a click chemistry-enabled, pre-targeting therapeutic and the first use of *in vivo* click chemistry in humans
▪ SQ3370 up to15x standard doxorubicin dose in patients with solid tumors was safe with no DLTs reported
▪ In Phase 2a, SQ3370 provided an unconfirmed objective response rate of 14.3% and disease control rate of 71.4% in patients with advanced sarcomas
▪ Tumor size reductions seen in both injected and non-injected lesions, potentially due to systemic anti-tumor responses

## INTRODUCTION

Only a small fraction (<1%) of cancer drugs reach the tumor, with the remaining dose eliminated by normal tissues.^1–3^ This can result in toxicities and adverse effects that limit the amount of drug that can be administered, which in turn compromises efficacy.^4–7^ Even widely used chemotherapies are often limited by their narrow therapeutic index, which is a measure of the effective dose of a drug relative to its safety. Additionally, there are a number of chemotherapeutic agents that are known to be efficacious in killing cancer cells but are too toxic for systemic administration, e.g. monomethyl auristatin E (MMAE).^8–11^

Antibody-drug conjugates (ADCs) have been developed to overcome the toxicity challenges associated with systemic drug administration; however off-target toxicities caused by catabolic activation hamper the dose of ADCs that can be given.^12,13^ About 99% of the dose of an ADC is catabolized by normal tissues that break the chemical linker between the antibody and the cytotoxic payload, leading to premature release of the active payload into circulation and off-target toxicities.^14,15^ Clinical data show that premature release of the active payload into circulation is not solved by stabilizing the linker.^16,17^ Other approaches for targeted delivery depend on other biological characteristics of tumors required for drug activation, such as oxygen level,^18,19^ pH,^20,21^ or enzymatic activity.^13,22–28^ As such, there is still a need to develop safe and effective cancer therapies that can be activated directly at tumor sites.

Pre-targeting is an approach that has been explored in the radiopharmaceutical industry for over a decade.^29,30^ Using this approach, the tumor targeted vehicle is uncoupled from the cytotoxic agent; each is administered separately and then reunited at the tumor. This can limit exposure to normal cells and reduce off-target toxicities.^29,30^

Shasqi, Inc. (San Francisco, CA, USA) developed a pre-targeting approach termed the Click Activated Protodrugs Against Cancer (CAPAC™) platform to increase the therapeutic index of cancer drugs by activating them at the tumor, thereby reducing systemic toxicities. This is achieved through the use of *in vivo* click chemistry at the site of the tumor. Click chemistry is a simple, powerful, Nobel prize-winning technology^31^ in which two molecules react only with each other, with no interference from or to the native biological processes.^32–35^ It is an ideal technology for pre-targeting as the reaction is rapid, covalent, selective *in vivo*, and nonimmunogenic.

The CAPAC platform consists of a tetrazine-containing clickable pre-targeting agent and a trans-cyclooctene-containing chemically attenuated clickable cancer drug (protodrug) administered separately and sequentially.^36–38^ These components click together at the site of the tumor through an irreversible covalent reaction between the tetrazine and *trans*-cyclooctene moieties, thereby releasing the active drug (or payload) into the tumor microenvironment.^36–38^ This pre-targeting approach eliminates catabolic activation by normal tissues, maximizing active drug concentration in the tumor while simultaneously reducing efficacy-limiting systemic toxicities.

SQ3370 is the first-in-class click-activated, pre-targeting therapy. It consists of: 1) an intratumorally injected clickable biopolymer (SQL70), which is a derivative of sodium hyaluronate, chemically modified with tetrazine, and 2) a protodrug (SQP33), which consists of a Dox payload whose cytotoxic activity is chemically attenuated by modification with *trans*-cyclooctene.^37,38^ When the SQP33 protodrug meets the SQL70 biopolymer, a rapid click reaction occurs between the tetrazine moiety of SQL70 and the *trans*-cyclooctene moiety of SQP33 (the click chemistry groups with complementary reactivity), selectively releasing Dox at the tumor and the tumor microenvironment.

Soft tissue sarcomas (STSs) are rare heterogeneous solid tumors of connective tissues that arise from mesenchymal precursors, most often in the extremities, and account for about 1% of adult cancers and 15% of childhood malignancies.^39–42^ Standard first-line systemic therapy for advanced STS disease includes Dox alone or in combination with other anti-cancer therapies.^42^ First-line Phase 3 trials comparing Dox monotherapy vs gemcitabine plus docetaxel,^43^ Dox plus ifosfamide,^44^ Dox plus palifosfamide,^45^ Dox plus evofosfamide,^19^ or Dox plus olaratumab^19^ did not show improvement in median overall survival of about 14 to 20 months and 2-year survival rates of 20% to 30%.

We conducted a Phase 1/2a open-label study to evaluate safety and efficacy of SQ3370 in patients with locally advanced or metastatic solid tumors. Here we report on the drug exposure, pharmacokinetics, recommended Phase 2 dose (RP2D), safety, and efficacy of SQ3370 in Phase 1 and on the safety, pharmacokinetics, efficacy, and immune effects of SQ3370 in Phase 2a.

## MATERIALS AND METHODS

### Patients

This was a first-in-human, multicenter, Phase 1/2a, single-arm, open-label, dose escalation and dose expansion study to evaluate the safety, tolerability, pharmacokinetics, and efficacy of SQ3370 in patients with locally advanced or metastatic solid tumors (SQ3370-001; NCT04106492). The study was conducted at 11 centers in the United States and Australia. Key inclusion criteria for the overall study were ≥ 18 years of age, pretreated histologically or cytologically confirmed cancer or desmoid tumor; an injectable tumor (confirmed by the Response Evaluation Criteria in Solid Tumors version 1.1 [RECIST v1.1], palpable or able to be injected percutaneously, and was accessible for repeated intratumoral or peritumoral injection with an 18- to 22-gauge needle), Eastern Cooperative Oncology Group (ECOG) performance status of 0 to 1, and adequate organ function. Key exclusion criteria were prior lifetime exposure to >300 mg/m^2^ of Dox HCl or DOXIL/CAELYX^®^ or 600 mg/m^2^ of epirubicin HCl or 600 mg/m² daunorubicin prior to study dosing; congestive heart failure, severe myocardial insufficiency, or cardiac arrhythmia; and major surgery, radiotherapy, or chemotherapy within 28 days prior to cycle 1 day 1. An additional inclusion criterion for patients enrolled in Phase 2a was the presence of metastatic STS with predefined anthracycline-sensitive histologies and no prior anthracycline treatment.

The study protocol was approved by an Institutional Review Board or Independent Ethics Committee at each participating site. The study was conducted in accordance with the principles of the Declaration of Helsinki, the Good Clinical Practice guidelines of the International Conference on Harmonization, and applicable regulatory requirements. Written informed consent was obtained from all study participants before any study procedures were conducted.

The study was overseen by a Safety Review Committee (SRC) led by the Medical Monitor and investigators. The SRC reviewed all available safety data at the completion of cycle 1 in Phase 1 for each cohort to evaluate possible dose-limiting toxicities (DLTs), recommend whether more patients were to be enrolled at a given dose level/cohort, and whether to dose escalate or stop enrollment based on occurrence of DLTs in addition to the SRC members’ best clinical judgment and provided recommendations to the sponsor. Phase 2a had predefined evaluations based on group enrollment, safety, and clinical activity.

### SQ3370 administration

SQ3370 consists of 2 components: the SQL70 clickable biopolymer and the SQP33 Dox protodrug. Details of drug administration are provided in Supplementary Methods and Supplementary Fig S1. Briefly, SQL70 biopolymer was administered intra and/or peritumorally on day 1 of each 21-day cycle into a lesion or lesions at a fixed 10 mL or 20 mL injection volume, followed by 3 or 5 consecutive daily infusions of SQP33 protodrug (day 1 to day 3 or day 5) with the cycle repeated 21 days after initiation of the previous cycle. SQ3370 doses were designed to deliver different Dox equivalents (Dox Eq) of the standard Dox dose, calculated as 1 g of SQP33 protodrug is equivalent to 0.7153 g of doxorubicin hydrochloride (1x). The Phase 1 Dose Escalation received SQ3370 administered at 0.38x to 15x Dox Eq dose (Dose Escalation Group 1) with 10 ml of biopolymer, while the Phase 1 Dose Escalation Group 2 received SQ3370 administered at 4x to 6x Dox Eq dose (Dose Escalation Group 2) with 20 ml of biopolymer, the Phase 2a Dose Expansion Extremity Soft-Tissue Sarcoma (STS) Group 1 received SQ3370 administered at the RP2D of 12X Dox Eq and 10 ml of biopolymer (Extremity STS Group 1), and the Phase 2a Dose Expansion Unresectable STS Group 2 received SQ3370 administered at the RP2D of 12X Dox over 3 vs 5 days and 20 ml of biopolymer (Unresectable STS Group 2). No patients were enrolled in the Phase 2a Expansion Head and Neck Group 3 cohort.

### Study endpoints

For both Phase 1 and Phase 2a, the primary endpoint was safety, including frequency of treatment emergent adverse events (TEAEs), serious TEAEs, DLTs, changes from baseline in vital signs, clinical laboratory parameters, physical examination findings, and ECG and ECHO/MUGA results. Secondary endpoints for both phases were determining plasma concentration and pharmacokinetic parameters for SQP33 protodrug and active Dox, and objective response rate (ORR) per RECIST v1.1. An additional primary endpoint for the Phase 1 portion was the determination of the RP2D. Additional secondary endpoints for Phase 2a were overall survival (OS) and time from enrollment to the first subsequent therapy. Additional key secondary endpoints for the Extremity STS Group 1 cohort included histopathologic response, disease-free survival, and local recurrence-free survival. For the Unresectable STS Group 2, additional key secondary endpoints included comparing the safety, tolerability, and pharmacokinetics of the 3-day vs 5-day dosing schedules of SQP33 protodrug, ORR ≥30%, and progression-free survival (PFS). For both Phase 1 and 2a, exploratory endpoints assessed the presence of SQP33 protodrug and active Dox in tumor tissue and the characterization of immune response in blood and tumor tissue over time following SQ3370 treatment. Details of study assessments are provided in Supplementary Methods.

### Statistical analysis

Details for sample size selection are summarized in Supplementary Methods. Statistical analysis methods for the safety and efficacy data were descriptive in nature. Continuous variables were summarized using descriptive statistics, including number of non-missing observations, mean, standard deviation, median, and minimum and maximum values. Categorical variables were summarized with frequency counts and percentages. Time-to-event variables were summarized using Kaplan-Meier (KM) methods. The safety analysis set included all patients who received at least one dose of SQL70 biopolymer and/or SQP33 protodrug. The efficacy analysis set included all patients who received at least 1 dose of SQL70 biopolymer and 1 dose of SQP33 protodrug and had at least 1 post-baseline tumor evaluation. Immune data were analyzed using Prism v10 (GraphPad Software, Boston, MA, USA). *P* < 0.05 was defined as statistically significant. *P*-values of cytometry by time of flight (CyTOF) data were obtained using two-way analysis of variance (ANOVA) with *post-hoc* Dunnett’s test (for comparison of immune contexture) and mixed-effects analysis with Geisser-Greenhouse correction and *post-hoc* Tukey’s test (for comparison of subpopulations). *P*-values of multiplex immunohistochemistry data were obtained by Mann-Whitney test (for comparisons between baseline and after first cycle) and by Log-rank Mantel-Cox analysis with Log-rank Hazard Ratio and 95% confidence intervals (for KM curve).

### Study termination

As adequate data had been generated to demonstrate clinical proof of concept of the click chemistry reaction in vivo, the sponsor recommended termination of the study following the first planned interim analysis, a decision that was supported by the SRC. The study was terminated on 07 September 2023. For the Phase 1 cohorts, data reported in this article are from the protocol-specified interim analysis with the data cut-off date of 15 December 2022, with no further analyses performed up to the study termination date. For the Phase 2a cohorts, data reported in this article are up to the study termination date of 07 September 2023.

## RESULTS

### Patients

Treatment and study discontinuations for the Phase 1 and 2a cohorts are presented in Supplementary Fig S2, Table 1, and Supplementary Tables S1 and S2. Phase 1 enrolled 38 patients; 30 patients in the Dose Escalation Group 1 cohort who received ≥1 dose of 10 mL SQL70 biopolymer and SQP33 protodrug at 0.38x to 15x Dox Eq dose and 8 in the Dose Escalation Group 2 cohort who received ≥1 dose of 20 mL SQL70 biopolymer and SQP33 protodrug at 4x or 6x Dox Eq dose (Supplementary Fig S2A, Supplementary Tables S1 and S2). The most common reason for treatment discontinuation was disease progression; all patients were terminated from the study due to the decision of the sponsor. One patient in Phase 1 received 8.8x Dox Eq dose.

**Table 1:**
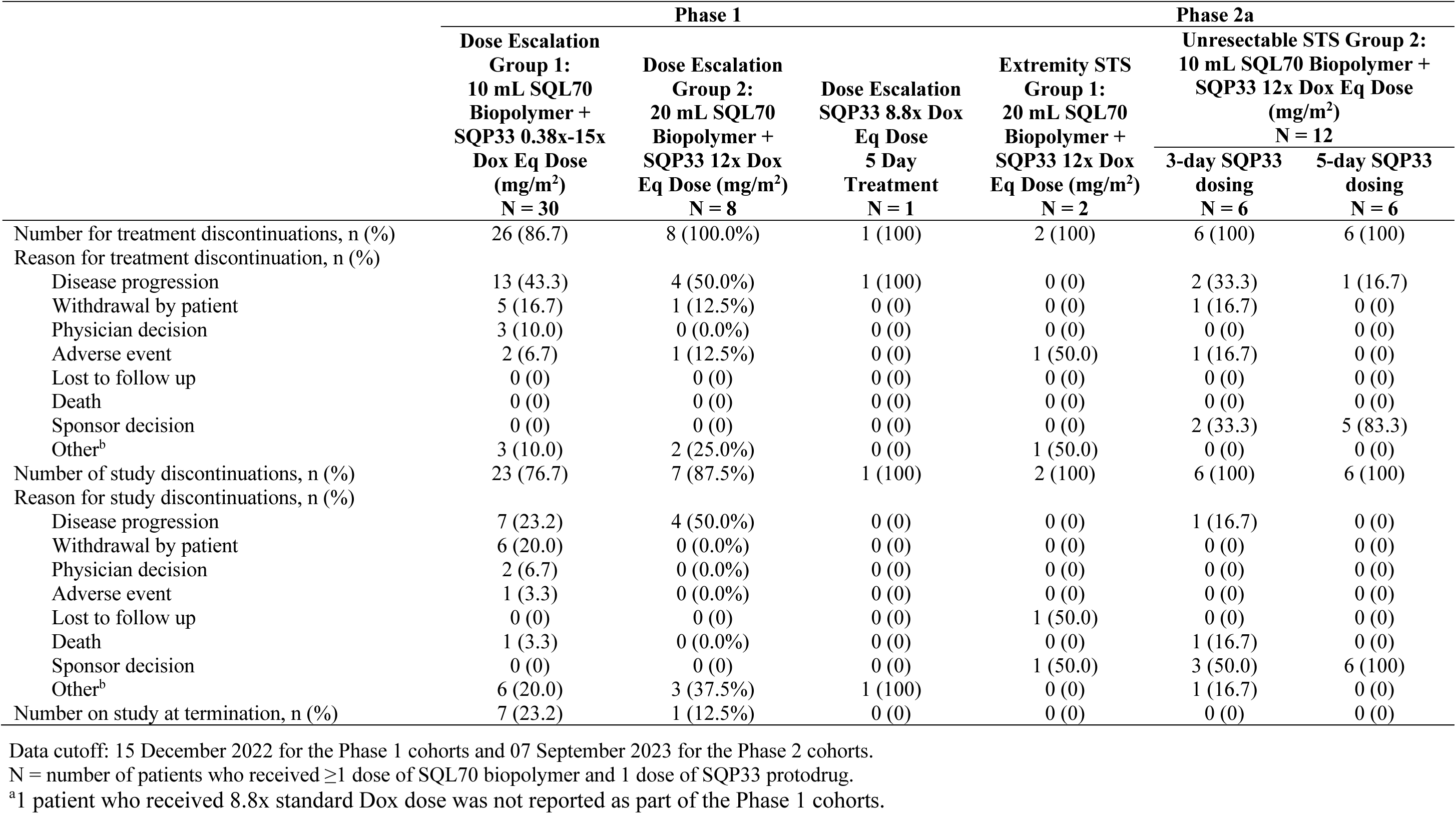
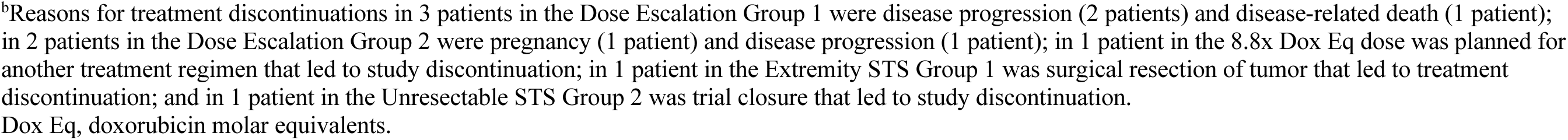
Patient disposition.

Phase 2a enrolled 14 patients before study termination; 2 patients in the Extremity STS Group 1 cohort and 12 in the Unresectable STS Group 2 cohort (6 patients in each of the 3-day and 5-day SQP33 dosing groups). The 2 patients in Group 1 received 20 mL SQL70 biopolymer and SQP33 protodrug while all the 12 patients in Group 2 received ≥1 dose of 10 mL SQL70 biopolymer and SQP33 protodrug (Supplementary Fig S2B, Table 1). The most common reason for study withdrawal was sponsor decision to end treatment and end study (10/14 [71.4%) patients, with 2/14 (14.3%) patients having withdrawn due to a TEAE before study termination.

### Baseline characteristics

The enrolled Phase 1 cohort patients (n=39) were 56.4% female, 87.2% white (Table 2). Patients had predominantly metastatic disease (77.9%); 87.4% had prior surgery, 48.7% prior radiotherapy, and 35.6% had previously received an immune checkpoint inhibitor. The enrolled Phase 2a cohort patients (n=14) were 50.0% female, 78.6% white. Patients had predominantly metastatic disease (78.6%); 85.6% had prior surgery, 50.0% prior radiotherapy, and 21.4% had previously received an immune checkpoint inhibitor.

**Table 2.**
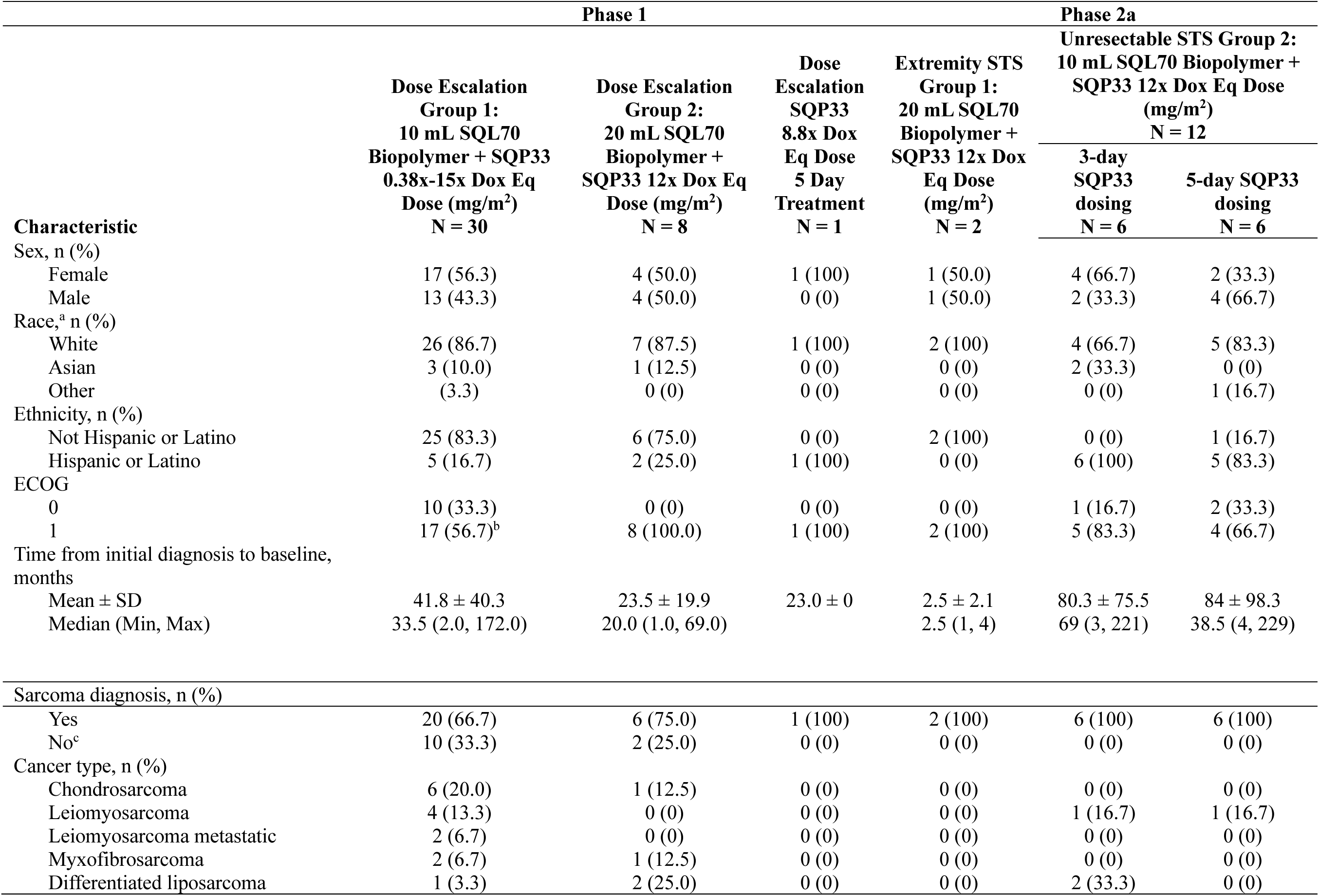
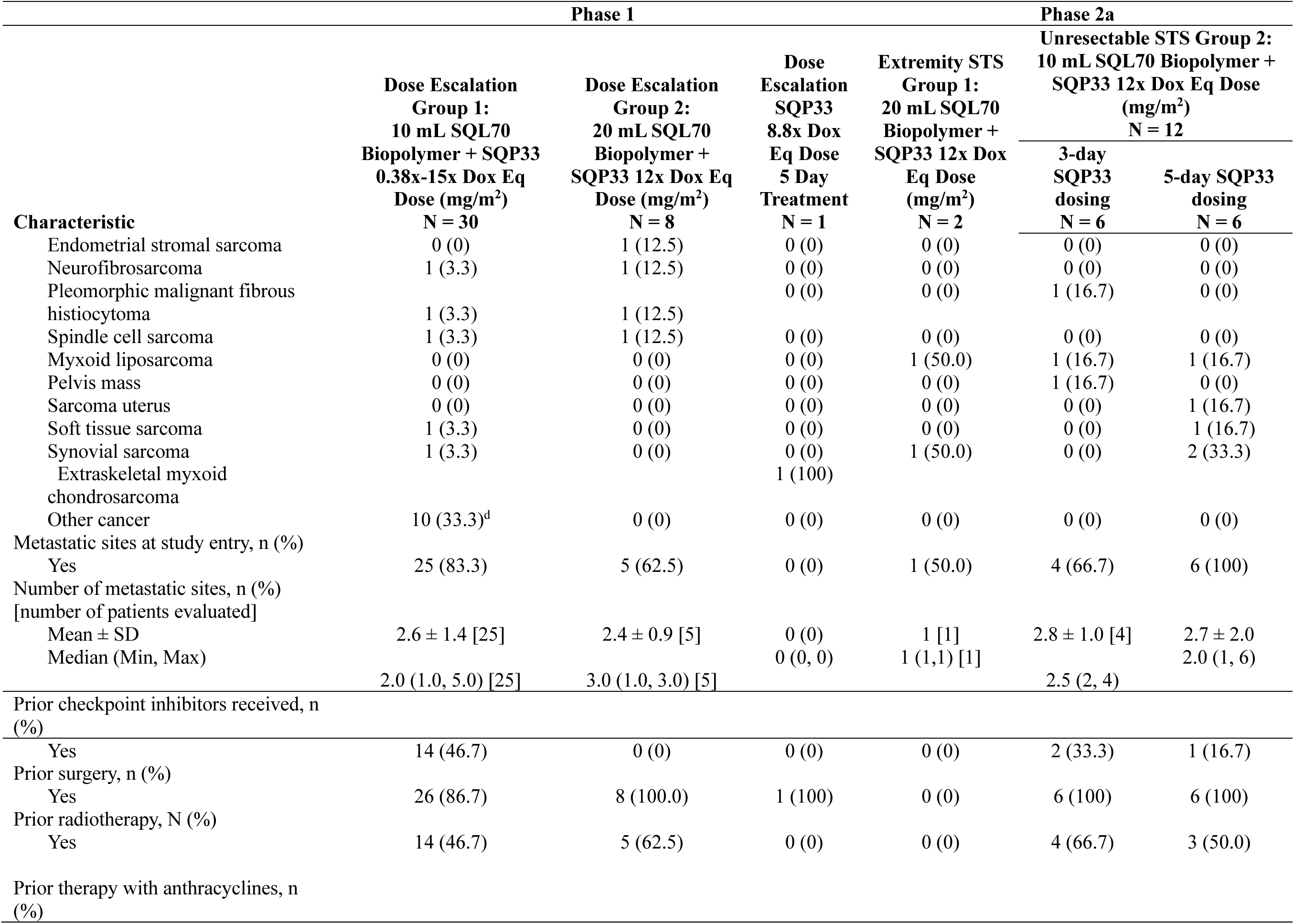
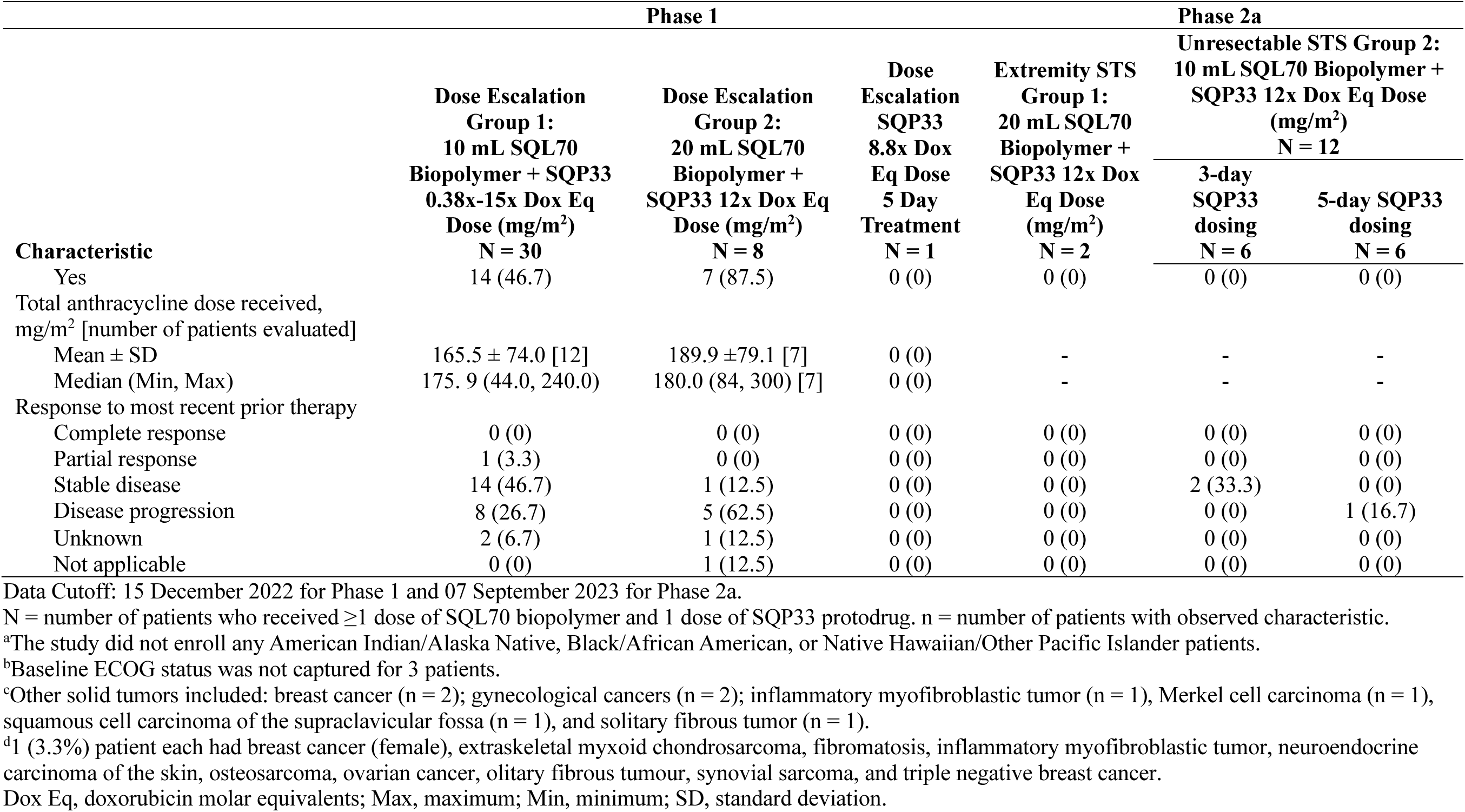
Baseline characteristics (safety population)

### SQ3370 exposure

SQ3370 exposure is provided in Table 3 and Supplementary Tables S3 and S4. Across the Phase 1 and 2a patients (n=53), more than 50% (54.7%; 29/53) received at least 4 treatment cycles of SQ3370 at the various Dox Eq doses; 15.5% (8/53) received at least 6 cycles, 5.7% (3/53) received at least 10 cycles, and 1.9% (1/53) received 12 cycles (Table 3). For patients who received the 12x Dox Eq dose in Phase 2a, more than 50% (57.1; 8/14) received at least 6 treatment cycles of SQ3370.

**Table 3:** SQ3370 administration and exposure.

|  | Phase 1 |  |  |  | Phase 2a |  |
| --- | --- | --- | --- | --- | --- | --- |
|  | Dose Escalation<br>Group 1:<br>10 mL SQL70<br>Biopolymer + SQP33<br>0.38x-15x Dox Eq<br>Dose (mg/m<sup>2</sup>)<br>N = 30 | Dose Escalation<br>Group 2:<br>20 mL SQL70<br>Biopolymer +<br>SQP33 12x Dox Eq<br>Dose (mg/m<sup>2</sup>)<br>N = 8 | Dose Escalation<br>SQP33 8.8x Dox<br>Eq Dose<br>5 Day<br>Treatment<br>N = 1 | Extremity STS<br>Group 1:<br>20 mL SQL70<br>Biopolymer +<br>SQP33 12x Dox<br>Eq Dose (mg/m<sup>2</sup>)<br>N = 2 | Unresectable STS Group 2:<br>10 mL SQL70 Biopolymer + SQP33<br>12x Dox Eq Dose (mg/m<sup>2</sup>)<br>N = 12 |  |
|  |  |  |  |  | 3-day SQP33<br>dosing<br>N = 6 | 5-day SQP33<br>dosing<br>N = 6 |
| Time on study, days |  |  |  |  |  |  |
| Mean ± SD | 109.1 ± 96.6 | 77.0 ± 41.3 | 95 ± 0 | 168.6 ± 34.7 | 110.3 ± 74.5 | 122 ± 74.5 |
| Median (Min, Max) | 67.5 (16.0, 393.0) | 67.5 (29.0, 144.0) | 95 (95) | 168.5 (144.0, 193.0) | 99.5 (41.0, 241.0) | 103.5 (50.0, 236.0) |
| Duration of SQP3370 therapy, days |  |  |  |  |  |  |
| Mean ± SD | 76.3 ± 80.7 | 45.1 ± 34.6 | 68 | 75.0 ± 69.3 | 63.8 ± 36.1 | 92.3 (72.7) |
| Median (Min, Max) | 49.9 (2, 362) | 36.0 (5.0, 117.0) | 68 (68, 68) | 75.0 (26.0, 124.0) | 66.5 (23.0, 108.0) | 71.5 (26.0, 207.0) |
| Number of SQL70 cycles started, n (%) |  |  |  |  |  |  |
| 1 | 3 (10.0) | 1 (12.5) | — | — | — | — |
| 2 | 11 (36.7) | 3 (37.5) | — | 1 (50.0) | 2 (33.3) | 5 (33.3) |
| 3 | 4 (13.3) | 2 (25.0) | 1 (100) | 0 (0) | 1 (16.7) | 2 (13.3) |
| 4 | 4 (13.3) | 1 (12.5) | — | 0 (0) | 1 (16.7) | 3 (20.0) |
| 5 | 1 (3.3) | — | — | 0 (0) | 1 (16.7) | 1 (16.7) |
| 6 | 3 (10.0) | 1 (12.5) | — | 1 (50.0) | 1 (16.7) | 2 (13.3) |
| 7 | 1 (3.3) | — | — | 0 (0) | 0 (0) | 0 (0) |
| 8 | 1 (3.3) | — | — | 0 (0) | 1 (16.7) | 1 (16.7) |
| 9 | 1 (3.3) | — | — | 0 (0) | 0 (0) | 0 (0) |
| 10 | 1 (3.3) | — | — | 0 (0) | 1 (16.7) | 1 (16.7) |
| 12 | 1 (3.3) | — | — | — | — | — |
| 18 | — | — | — | — | — | — |
| SQP33 cumulative dose, mg/m<sup>2</sup> |  |  |  |  |  |  |
| Mean ± SD | 2939.7 ± 2421.3 | 1367.3 ± 1015.5 | 2590.6 | 4999.3 ± 3528.1 | 4534.1 ± 2096.1 | 5950.4 ± 3719.3 |
| Median (Min, Max) | 2381.2 (162.0, 11000.0) | 1062.7 (626.0, 3726.0) | 2590.6 (2590.6, 2590.6) | 4999.3 (2505.0, 7494.0) | 4376.1 (2252.0, 7492.0) | 4947.0 (2490.1, 11760.0) |
| SQL70 dose interruptions |  |  |  |  |  |  |
| No | 30 (100.0) | 8 (100.0) | 1 (100) | 2 (100) | 6 (100) | 6 (100) |
| Yes | 0 (0.0) | 0 (0) | 0 (0) | 0 (0) | 0 (0) | 0 (0) |
| SQL70 dose modifications |  |  |  |  |  |  |
|  | Dose Escalation<br>Group 1:<br>10 mL SQL70<br>Biopolymer + SQP33<br>0.38x-15x Dox Eq<br>Dose (mg/m<sup>2</sup>)<br>N = 30 | Dose Escalation<br>Group 2:<br>20 mL SQL70<br>Biopolymer +<br>SQP33 12x Dox Eq<br>Dose (mg/m<sup>2</sup>)<br>N = 8 | Dose Escalation<br>SQP33 8.8x Dox<br>Eq Dose<br>5 Day<br>Treatment<br>N = 1 | Extremity STS<br>Group 1:<br>20 mL SQL70<br>Biopolymer +<br>SQP33 12x Dox<br>Eq Dose (mg/m<sup>2</sup>)<br>N = 2 | Unresectable STS Group 2:<br>10 mL SQL70 Biopolymer + SQP33<br>12x Dox Eq Dose (mg/m<sup>2</sup>)<br>N = 12 |  |
|  |  |  |  |  | 3-day SQP33<br>dosing<br>N = 6 | 5-day SQP33<br>dosing<br>N = 6 |
| No | 29 (96.7) | 7 (87.5) | 1 (100) | 2 (100) | 6 (100) | 6 (100) |
| Yes | 1 (3.3) | 1 (12.5) | 0 (0) | 0 (0) | 0 (0) | 0 (0) |
| Decrease in dose | 0 (0) | 0 (0) | 0 (0) | 0 (0) | 0 (0) | 0 (0) |
| SQP33 dose interruptions |  |  |  |  |  |  |
| No | 29 (96.7) | 8 (100.0) | 1 (100) | 2 (100) | 6 (100) | 6 (100) |
| Yes | 1 (3.3) | 0 (0) | 0 (0) | 0 (0) | 0 (0) | 0 (0) |
| SQP33 dose modifications |  |  |  |  |  |  |
| No | 28 (93.3) | 8 (100.0) | 1 (100) | 2 (100) | 5 (83.3) | 6 (100) |
| Yes | 2 (6.7) | 0 (0) | 0 (0) | 0 (0) | 0 (0) | 0 (0) |
| Decrease in dose | 0 (0) | 0 (0) | 0 (0) | 0 (0) | 1 (16.7) | 0 (0) |

Data Cutoff: 15 December 2022 for the Phase 1 cohorts and 07 September 2023 for the Phase 2 cohorts.
N = number of patients who received $\geq 1$ dose of SQL70 biopolymer and 1 dose of SQP33 protodrug.
Dox Eq, doxorubicin molar equivalents; Max, maximum; Min, minimum; SD, standard deviation.

In the Phase 1 Dose Escalation Group 1 cohort, median (range) time on SQP3370 therapy was 49.9 (2, 362) days and median (range) SQP33 cumulative dose was 2381.1 (162.0, 11000.0) mg/m^2^ (Table 3). In the Phase 1 Dose Escalation Group 2 cohort, median (range) time on SQP3370 therapy was 36 (5.0, 117) days and median (range) SQP33 cumulative dose was 1062.7 (626.0, 3726.0) mg/m^2^. The patient who received 8.8x Dox Eq dose received SQP3370 therapy for 68 days and a cumulative dose of SQP33 of 2590.6 mg/m^2^. At study termination, in the Phase 2a Extremity STS Group 1 cohort, median (range) time on SQP3370 therapy and median (range) SQP33 cumulative dose were 75 (26, 124) days and 4999.3 (2505.0, 7494.0) mg/m^2^, respectively; in the Phase 2a Unresectable STS Group 2 cohort with the 3-day SPQ33 dosing schedule were 67 (23, 108) days and 4376.1 (2252.0, 7492.0) mg/m^2^, respectively; and in the Phase 2a Unresectable STS Group 2 cohort with the 5-day SPQ33 dosing schedule were 72 (26, 207) days and 4947.0 (2490.1, 11760.0) mg/m^2^, respectively.

### SQP33 and Dox pharmacokinetics following SQ3370 treatment

Plasma pharmacokinetics data of patients treated with SQ3370 in Phase 1 have been previously reported.^46^ On cycle 1 day 1, infused SQP33 was rapidly cleared from the plasma in the presence of SQL70 biopolymer, and this was associated with an increase in circulating plasma Dox. No further increase in the Dox concentration with increasing doses of SQ3370 supported the selection of 12x as the RP2D (Supplementary Table S5). Similar pharmacokinetic profiles were observed between the Phase 1 12x Dox Eq dose cohort and the Phase 2a Group 1 and Group 2 cohorts that had also received the 12x Dox Eq dose (Fig 1). There were changes in Phase 2a Group 2 with a 3-day dosing regimen, where the patients were administered SQP33 at a Dox dose of 250 mg/m^2^ on day 1 and 500 mg/m^2^ on days 2 and 3 (Supplementary Table S5). Pharmacokinetic parameters for 3-day vs 5-day dosing schedules of SQ3370 showed that the mean profile of SQP33 was higher on day 3 in the 3-day cohort than in the 5-day cohort, but still with similar dox exposure. The doxorubicin AUC_last_ for 3-day and 5-day were calculated as 3886.8 h*ng/mL and 2368.5 h*ng/mL, respectively.

**Fig 1.**
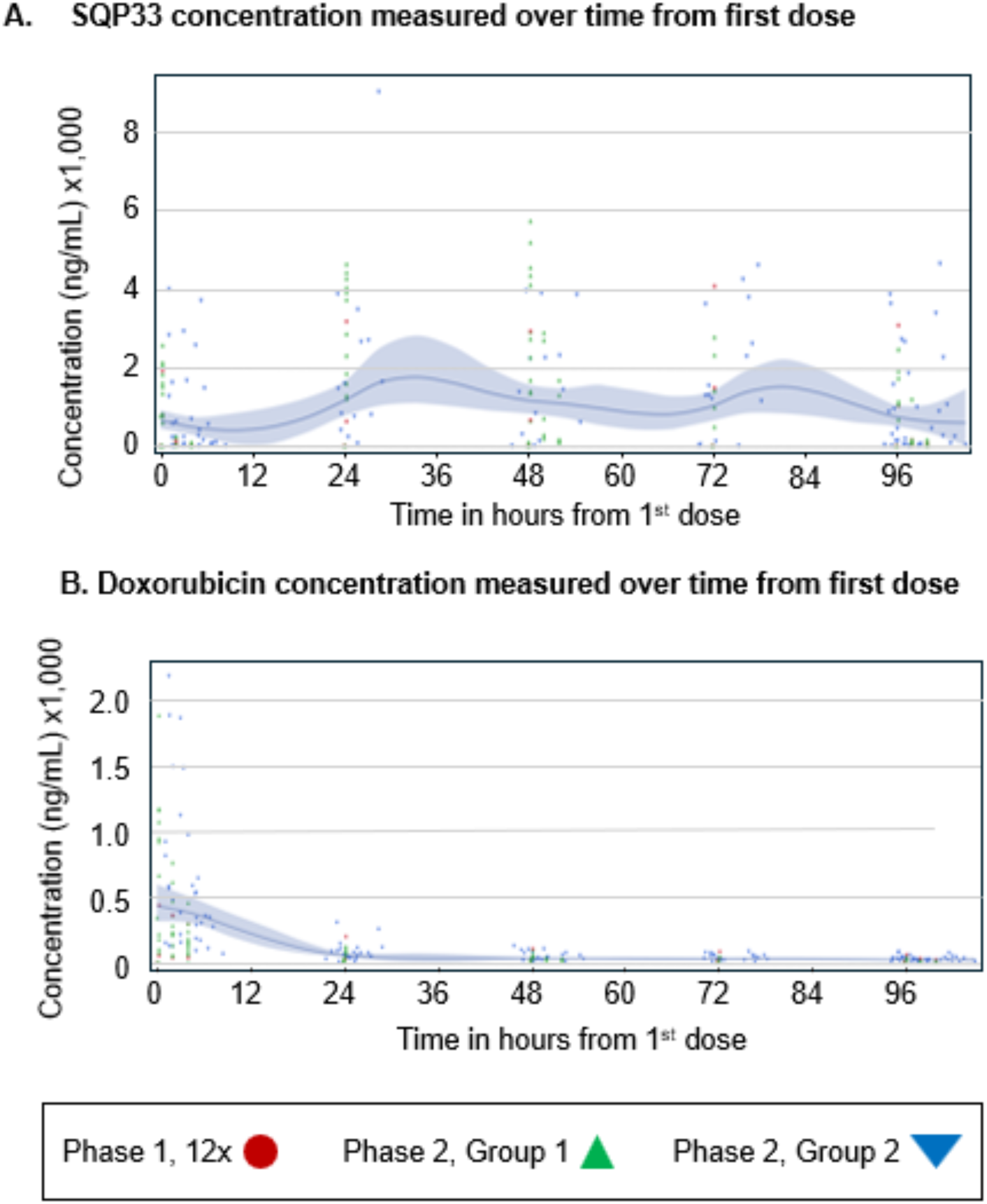
SQ3370 pharmacokinetics in Phase 1/2a patients for A) SQP33 concentration and B) Dox concentration over time. Shown are values for individual patients, labeled based on their respective cohort. Solid lines represent the average concentrations, and the grey areas indicate the ± standard deviation. Dox, doxorubicin.

### TEAEs, serious TEAEs, and DLTs

A total of 290 TEAEs were reported in the Phase 1 Dose Escalation Group 1 cohort, with each of the 30 (100.0%) patients who received SQ3370 reporting ≥1 TEAE (Table 4, Supplementary Table S6). The most common TEAEs were nausea in 17 (56.7%) patients, fatigue in 15 (50.0%), and constipation in 9 (30.0%) patients. TEAEs related to study drug were reported in 24 (80.0%) patients and serious TEAEs were reported in 10 (33.3%) patients (Supplementary Table S6). TEAEs leading to study discontinuation were reported in 3 (10.0%) patients, with 1 (3.3%) patient each discontinuing the study due to transient cardiac failure (unrelated and reversable), fatigue, and dyspnea (Table 4, Supplementary Table S6). DLTs were not reported in any patients, and Grade 3 or 4 TEAEs were reported in 12 (40.0%) patients. One (3.3%) death occurred in the 1 patient who received SQ3370 at 0.38x Dox Eq dose. The patient died of a thrombotic stroke that became hemorrhagic after thrombolysis; the stroke was deemed unrelated to the study drug by the investigator. Mild anemia, neutropenia, and thrombocytopenia were reported across the SQ3370 doses (Table 4).

**Table 4:** TEAEs in Phase 1 and Phase 2a cohorts treated with SQ3370.

| TEAE Type | Phase 1 |  |  |  | Phase 2a |  |
| --- | --- | --- | --- | --- | --- | --- |
|  | Dose Escalation<br>Group 1:<br>10 mL SQL70<br>Biopolymer + SQP33<br>0.38x-15x Dox Eq<br>Dose (mg/m<sup>2</sup>)<br>N = 30 | Dose Escalation<br>Group 2:<br>20 mL SQL70<br>Biopolymer +<br>SQP33 12x Dox Eq<br>Dose (mg/m<sup>2</sup>)<br>N = 8 | Dose Escalation<br>SQP33 8.8x Dox<br>Eq Dose<br>5 Day Treatment<br>N = 1 | Extremity STS<br>Group 1:<br>20 mL SQL70<br>Biopolymer +<br>SQP33 12x Dox<br>Eq Dose (mg/m<sup>2</sup>)<br>N = 2 | Unresectable STS Group 2:<br>10 mL SQL70 Biopolymer +<br>SQP33 12x Dox Eq Dose<br>(mg/m<sup>2</sup>)<br>N = 12 |  |
|  |  |  |  |  | 3-day SQP33 | 5-day SQP33 |
|  |  |  |  |  | dosing<br>N = 6 | dosing<br>N = 6 |
| Number of TEAEs reported | 290 | 63 | 5 | 12 | 56 | 44 |
| Patients with TEAEs, n (%) | 30 (100.0) | 8 (100.0) | 1 (100) | 2 (100) | 6 (100) | 6 (100) |
| Any TEAEs | 30 (100.0) | 8 (100.0) | 1 (100) | 2 (100) | 6 (100) | 6 (100) |
| Most common TEAEs (in ≥ 20%<br>of patients), n (%) |  |  |  |  |  |  |
| Nausea | 17 (56.7) | 4 (50.0) | 0 (0) | 1 (50.0) | 5 (83.3) | 6 (100) |
| Fatigue | 15 (50.0) | 2 (25.0) | 0 (0) | 2 (100) | 4 (66.7) | 4 (66.7) |
| Constipation | 9 (30.0) | 0 (0) | 0 (0) | 0 (0) | 3 (50.0) | 1 (16.7) |
| Decreased appetite | 7 (23.3) | 4 (50.0) | 0 (0) | 1 (50.0) | 0 (0) | 1 (16.7) |
| Anemia | 7 (23.3) | 4 (50.0) | 0 (0) | 0 (0) | 0 (0) | 1 (16.7) |
| Neutropenia | 4 (13.3) | 2 (25.0) | 0 (0) | 1 (50.0) | 1 (16.7) | 2 (33.3) |
| Diarrhea | 3 (10.0) | 2 (25.0) | 0 (0) | 0 (0) | 1 (16.7) | 2 (33.3) |
| Dizziness | 2 (6.7) | 2 (25.0) | 0 (0) | 0 (0) | 0 (0) | 0 (0) |
| TEAEs related to study drug | 24 (80.0) | 6 (75.0) | 1 (100) | 2 (100) | 6 (100) | 6 (100) |
| Serious TEAE | 10 (33.3) | 2 (25.0) | 0 (0) | 0 (0) | 3 (33.3) | 1 (16.7) |
| TEAE leading to study<br>discontinuation | 3 (10.0) | 0 (0) | 0 (0) | 1 (50.0) | 1 (16.7) | 1 (16.7) |
| Cardiac failure | 1 (3.3) | 0 (0) | 0 (0) | 0 (0) | 0 (0) | 0 (0) |
| Fatigue | 1 (3.3) | 0 (0) | 0 (0) | 0 (0) | 0 (0) | 0 (0) |
| Dyspnoe | 1 (3.3) | 0 (0) | 0 (0) | 0 (0) | 0 (0) | 0 (0) |
| Patients with DLTs | 0 (0) | 0 (0) | 0 (0) | 0 (0) | 0 (0) | 0 (0) |
| Patients with Grade 3 or 4 TEAEs | 12 (40.0) | 4 (50.0) | 0 (0) | 1 (50.0) | 2 (33.3) | 1 (16.7) |
| Patients with TEAEs related to death | 1 (3.3) | 0 (0) | 0 (0) | 0 (0) | 1 (16.7) | 0 (0) |
| Pneumonia | 0 (0) | 0 (0) | 0 (0) | 0 (0) | 1 (16.7) | 0 (0) |
| Patients with adverse events of<br>myelosuppression of ≥ Grade 3<br>severity, n (%) |  |  |  |  |  |  |
|  | Dose Escalation<br>Group 1:<br>10 mL SQL70<br>Biopolymer + SQP33<br>0.38x-15x Dox Eq<br>Dose (mg/m<sup>2</sup>)<br>N = 30 | Dose Escalation<br>Group 2:<br>20 mL SQL70<br>Biopolymer +<br>SQP33 12x Dox Eq<br>Dose (mg/m<sup>2</sup>)<br>N = 8 | Dose Escalation<br>SQP33 8.8x Dox<br>Eq Dose<br>5 Day Treatment<br>N = 1 | Extremity STS<br>Group 1:<br>20 mL SQL70<br>Biopolymer +<br>SQP33 12x Dox<br>Eq Dose (mg/m<sup>2</sup>)<br>N = 2 | Unresectable STS Group 2:<br>10 mL SQL70 Biopolymer +<br>SQP33 12x Dox Eq Dose<br>(mg/m<sup>2</sup>)<br>N = 12 |  |
|  |  |  |  |  | 3-day SQP33 | 5-day SQP33 |
|  |  |  |  |  | dosing<br>N = 6 | dosing<br>N = 6 |
| Anemia |  |  |  |  |  |  |
| Grade 3 | 4 (13.3) | 3 (37.5) | 0 (0) | 0 (0) | 0 (0) | 0 (0) |
| Grade 4 | 1 (3.3) | 0 (0) | 0 (0) | 0 (0) | 0 (0) | 0 (0) |
| Grade 5 | 0 (0) | 0 (0) | 0 (0) | 0 (0) | 0 (0) | 0 (0) |
| Neutropenia |  |  |  |  |  |  |
| Grade 3 | 1 (3.3) | 1 (12.5) | 0 (0) | 1 (50.0) | 1 (16.7) | 1 (16.7) |
| Grade 4 | 2 (6.7) | 1 (12.5) | 0 (0) | 0 (0) | 0 (0) | 0 (0) |
| Grade 5 | 0 (0) | 0 (0) | 0 (0) | 0 (0) | 0 (0) | 0 (0) |
| Febrile neutropenia |  |  |  |  |  |  |
| Grade 3 | 0 (0) | 0 (0) | 0 (0) | 0 (0) | 1 (16.7) | 0 (0) |
| Grade 4 | 0 (0) | 0 (0) | 0 (0) | 0 (0) | 0 (0) | 0 (0) |
| Grade 5 | 0 (0) | 0 (0) | 0 (0) | 0 (0) | 0 (0) | 0 (0) |
| Thrombocytopenia |  |  |  |  |  |  |
| Grade 3 | 0 (0) | 1 (12.5) | 0 (0) | 0 (0) | 1 (16.7) | 0 (0) |
| Grade 4 | 2 (6.7) | 0 (0) | 0 (0) | 0 (0) | 0 (0) | 0 (0) |
| Grade 5 | 0 (0) | 0 (0) | 0 (0) | 0 (0) | 0 (0) | 0 (0) |

Data Cutoff: 15 December 2022 for Phase 1 and 07 September 2023 for Phase 2a.
N = number of patients who received ≥1 dose of SQL70 biopolymer and 1 dose of SQP33 protodrug.
TEAEs were coded using MedDRA Version 24.1
AESI; adverse event of special interest; DLT, dose-limiting toxicity; Dox Eq, doxorubicin molar equivalents; MedDRA, Medical Dictionary for Regulatory Activities; max, maximum; min, minimum; SD, standard deviation; TEAE, treatment emergent adverse event.

A total of 63 TEAEs were reported in the Phase 1 Dose Escalation Group 2 cohort, with each of the 8 (100.0%) patients who received SQ3370 reporting ≥1 TEAE (Table 4, Supplementary Table S7). The most common TEAEs were nausea, decreased appetite, and anemia in 4 (50.0%) patients each. TEAEs related to the study drug were reported in 6 (75.0%) patients, and serious TEAEs were reported in 2 (25.0%) patients. No TEAEs leading to study discontinuation or DLTs were reported in any of the 8 patients. No study related deaths were reported. Mild anemia, neutropenia, and thrombocytopenia were reported across the Dox Eq doses of SQ3370 (Table 4).

Five TEAEs were reported in the 1 patient who received the 8.8x standard Dox dose (Table 4). The patient experienced a TEAE related to the study drug but had no serious TEAEs.

A total of 12 TEAEs were reported in the 2 patients who received SQ3370 in the Phase 2a Extremity STS Group 1 cohort, with each patient reporting ≥1 TEAE (Table 4). The most common TEAE was nausea, which was reported in two (100%) patients. TEAEs related to the study drug were reported in the 2 (100%) patients, and TEAEs leading to study discontinuation were reported in 1 (50.0%) patient. Grade 3 or 4 TEAEs were reported in 1 (50.0%) patient. No deaths were reported.

In the Phase 2a Unresectable STS Group 2 cohort with the 3-day and 5-day dosing sequence of SQP33, a total of 56 and 44 TEAEs, respectively, were reported, with all 6 patients in each group reporting TEAEs (Table 4). The most common TEAES reported with 3-day dosing were nausea in 5 (83.3%) patients, fatigue in 4 (66.7%), and constipation in 3 (50.0%). The most common TEAEs with 5-day dosing were nausea in 6 (100%) patients and fatigue in 4 (66.7%). TEAEs related to the study drug were reported in 6 (100%) patients in each group; serious TEAES were reported in 3 (50.0%) patients with 3-day dosing and 1 (16.7%) patient with 5-day dosing. TEAEs leading to study discontinuation were not reported in any patients in these groups. Grade 3 or 4 TEAEs were reported in 2 (33.3%) patients with 3-day dosing and 1 (16.7%) with 5-day dosing. A death due to pneumonia was reported in 1 (16.7%) patient with 3-day dosing. Mild neutropenia and thrombocytopenia were reported in these groups (Table 4). One patient (16.7%) in the 3-day dosing group had Grade 3 febrile neutropenia.

### ECG and ECHO/MUGA results

Across the Phase 1 and Phase 2a cohorts, no significant ECG changes in the QTcF over subsequent cycles, nor a significant decrease in the ECHO/MUGA LVEF. There was 1 episode of subacute systolic cardiac failure in a patient in the Phase 1 Dose Escalation Group 1 cohort who was non-compliant with treatment for adrenal insufficiency and hypothyroidism; the patient recovered on resumption of medical therapy.

### Efficacy

In the Phase 1 Dose Escalation Group 1 cohort (n=24), ORR was 0%, stable disease was the best response in 58.3% (14/24 patients), and disease progression occurred in 41.7% (10/24 patients) (Table 5). Disease control rate (DCR) was 58.3% (14/24 patients; 95% CI: 36.3, 77.9) and KM median duration of stable disease was 3.1 (95% CI: 1.3, 4.9) months. KM median PFS was 3.1 (95% CI: 1.3, 4.9) months. Reductions in tumor size were seen in injected lesions in Group 1 cohort patients evaluated, ORR was 0%, stable disease was the best response with unconfirmed partial response reported in 1 (4.2%) patient (Fig 2A). In the Phase 1 Dose Escalation Group 2 cohort (n=7), ORR was 0%, stable disease was 71.4% (5/7 patients) and disease progression was 28.6% (4/7 patients) (Table 5). DCR was 71.4% (5/7 patients; 95% CI: 29.0, 96.3) and KM median duration of stable disease was 2.6 (95% CI: 1.2, 4.3) months. KM median PFS was 2.6 (95% CI: 1.2, 4.3) months. Reductions in tumor size were seen in injected lesions across the Group 2 cohort patients evaluated, with no partial response reported in any patient (Fig 2A). The 1 patient who received 8.8x Dox Eq dose in the Phase 1 expansion cohort had stable disease.

**Fig 2.**
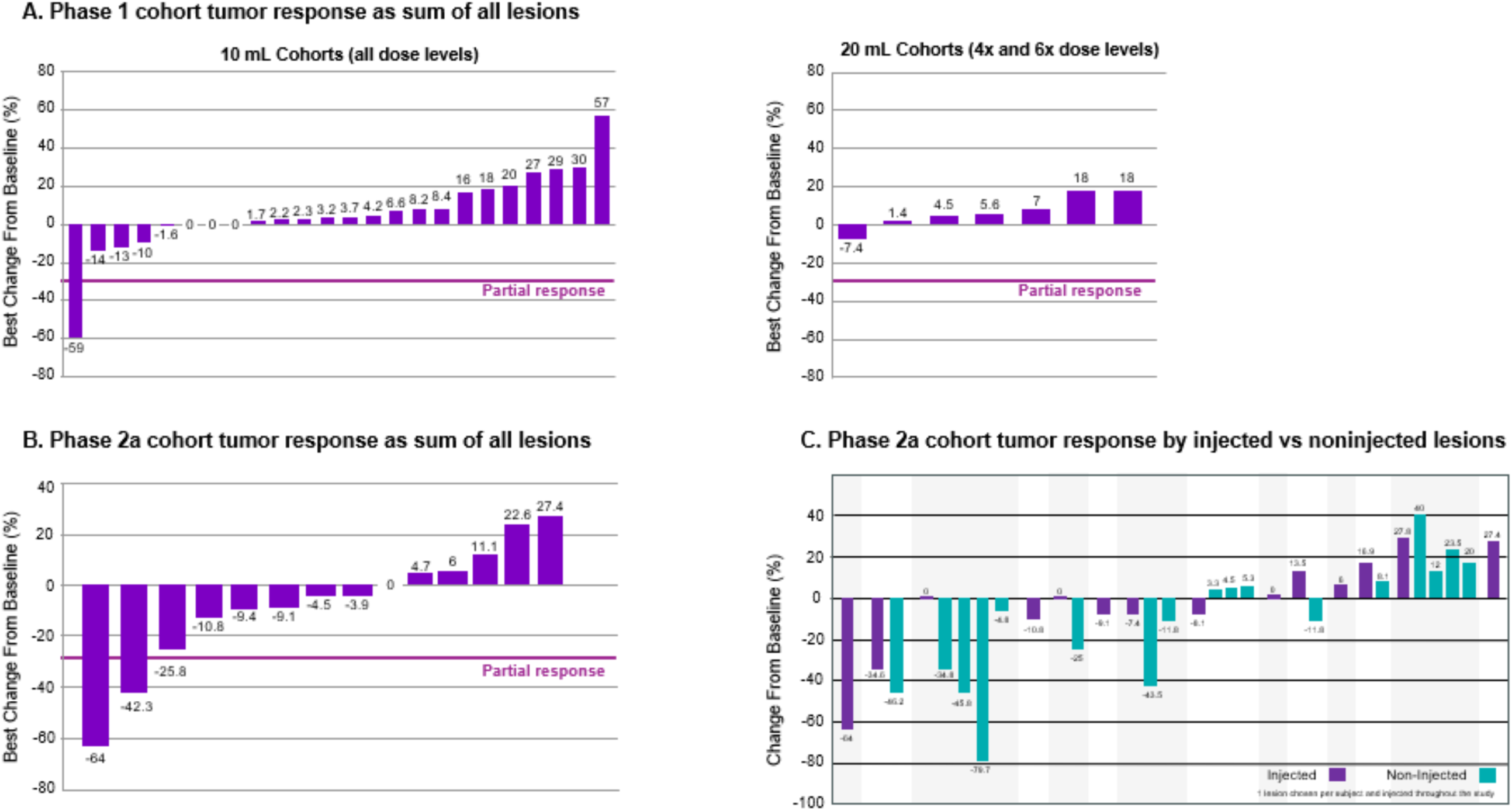
Tumor response in Phase 1/2a cohorts. A) Sum of all target lesions for Phase 1 patients who received SQP33 protodrug at different Dox Eq and either SQL70 biopolymer of 10 mL (n = 24) or 20 mL (n = 7) injection volume. B) Sum of all target lesions and C) injected vs non-injected lesions for Phase 2a patients (n = 14) who received SQP33 protodrug of 12x Dox Eq and 10 mL or 20 mL injection volume of SQL70 biopolymer. The efficacy set included all patients who received at least 1 dose of SQL70 biopolymer and 1 dose of SQP33 protodrug and had at least 1 post-baseline tumor evaluation. Dox Eq, doxorubicin equivalent.

**Table 5:** Efficacy in Phase 1 and Phase 2a cohorts treated with SQ3370.

| Outcome | Phase 1 |  |  | Phase 2a |  |  |  |
| --- | --- | --- | --- | --- | --- | --- | --- |
|  | Dose Escalation<br>Group 1:<br>10 mL SQL70<br>Biopolymer +<br>SQP33 0.38x-15x<br>Dox Eq Dose<br>(mg/m<sup>2</sup>)<br>N = 24 | Dose Escalation<br>Group 2:<br>20 mL SQL70<br>Biopolymer +<br>SQP33 12x Dox Eq<br>Dose (mg/m<sup>2</sup>)<br>N = 7 | Dose<br>Escalation<br>SQP33 8.8x<br>Dox Eq<br>5 Day<br>Treatment<br>N = 1 | Extremity<br>STS Group 1:<br>20 mL SQL70<br>Biopolymer +<br>SQP33 12x<br>Dox Eq Dose<br>(mg/m<sup>2</sup>)<br>N = 2 | Unresectable STS Group 2:<br>10 mL SQL70 Biopolymer +<br>SQP33 12x Dox Eq Dose (mg/m<sup>2</sup>)<br>3-day SQP33<br>dosing<br>N = 6 | 5-day SQP33<br>dosing<br>N = 6 | Total STS<br>Population: 10 or<br>20 mL SQL70<br>Biopolymer +<br>SQP33 12x Dox Eq<br>Dose (mg/m<sup>2</sup>)<br>N = 14 |
| Objective response rate, n (%) | 0 (0) | 0 (0) | 0 (0) | 1 (50.0) | 0 (0) | 1 (16.7) | 2 (14.3) |
| 95% exact CI | (0, 14.2) | (0, 41.0) | (0, 97.5) | (1.3, 98.7) | (0, 45.9) | (0.4, 64.1) | (1.8, 42.8) |
| Best overall response, n (%) |  |  |  |  |  |  |  |
| Complete response | 0 (0) | 0 (0) | 0 (0) | 0 (0) | 0 (0) | 0 (0) | 0 (0) |
| Partial response | 0 (0) | 0 (0) | 0 (0) | 1 (50.0) | 0 (0) | 1 (16.7) | 2 (43.3) |
| Stable disease | 14 (58.3) | 5 (71.4) | 1 (100) | 1 (50.0) | 4 (66.7) | 3 (50.0) | 8 (57.1) |
| Disease progression | 10 (41.7) | 2 (28.6) | 0 (0) | 0 (0) | 2 (33.3) | 2 (33.3) | 4 (28.6) |
| Not evaluable | 0 (0) | 0 (0) | 0 (0) | 0 (0) | 0 (0) | 0 (0) | 0 (0) |
| Disease control rate, n (%) | 14 (58.3) | 5 (71.4) | 1 (100) | 2 (100) | 4 (66.7) | 4 (66.7) | 10 (71.4) |
| 95% exact CI | (36.6, 77.9) | (29.0, 96.3) | (2.5, 100) | (15.8, 100) | (22.2, 95.7) | (22.3, 95.7) | (41.9, 91.6) |
| Duration of stable disease<br>(months) |  |  |  |  |  |  |  |
| KM median | 3.1 | 2.6 | — | — | — | — | — |
| 95% CI | (1.3, 4.9) | (1.2, 4.3) | — | — | — | — | — |
| (Q1, Q3) | (1.3, 6.8) | (1.2, 3.6) | — | — | — | — | — |
| (Min, Max) | (0.8, 12.9) | (0.7, 4.3) | — | — | — | — | — |

N = number of patients who received at least 1 dose of SQL70 biopolymer and 1 dose of SQP33 protodrug and had at least 1 post-baseline tumor evaluation.
CI, confidence interval; Dox Eq, doxorubicin molar equivalents; KM; Kaplan-Meier; Max, maximum; Min, minimum; ND, not determined. Q1, first quartile; Q3, third quartile.

At the time of study termination for Phase 2a STS cohort (n=14), ORR was 0%, unconfirmed ongoing ORR in 14.3% (2/14 patients), stable disease was best response for 57.1% (8/14 patients), disease progression was 28.6% (4/14 patients), and DCR was 71.4% (10/14 patients; 95% CI: 41.9, 91.6) (Table 5). As 5/14 (35.7%) patients had a PFS event by study termination, the KM median PFS could not be determined. No differences were observed between the 3-day and 5-day dosing SQP33 dosing schedules. Reductions in individual tumors were seen in both injected and non-injected lesions, with partial responses reported in 2 (14.3%) patients (Fig 2B, C).

Clinical activity was observed in 21.4% (3/14) of patients in the Phase 2a STS cohort. Patient 1 was a female with unresectable synovial sarcoma who was treated with SQ3370 for 2 cycles and achieved stable disease as shown on RECIST 1.1 with a cold PET scan (Fig 3A). However, this patient refused surgical amputation and was lost to follow-up. Patient 2 was a female with right chest-wall refractory metastatic undifferentiated pleomorphic sarcoma who started fifth-line therapy with SQ3370 for 4 cycles and achieved unconfirmed partial response (Fig 3B) at study termination. The patient had received prior first line therapy with gemcitabine/docetaxel, second line therapy with the PD-1 inhibitor envasaarc, third line therapy with the immunotherapies ipilimumab/nivolumab, and fourth line therapy with cabozantinib. Patient 3 was a male with metastatic liposarcoma whose primary tumor converted from unresectable to “resectable” after treatment with SQ3370 for 6 cycles, leading to complete surgical resection with negative tumor margins and 25% necrosis.

**Fig 3.**
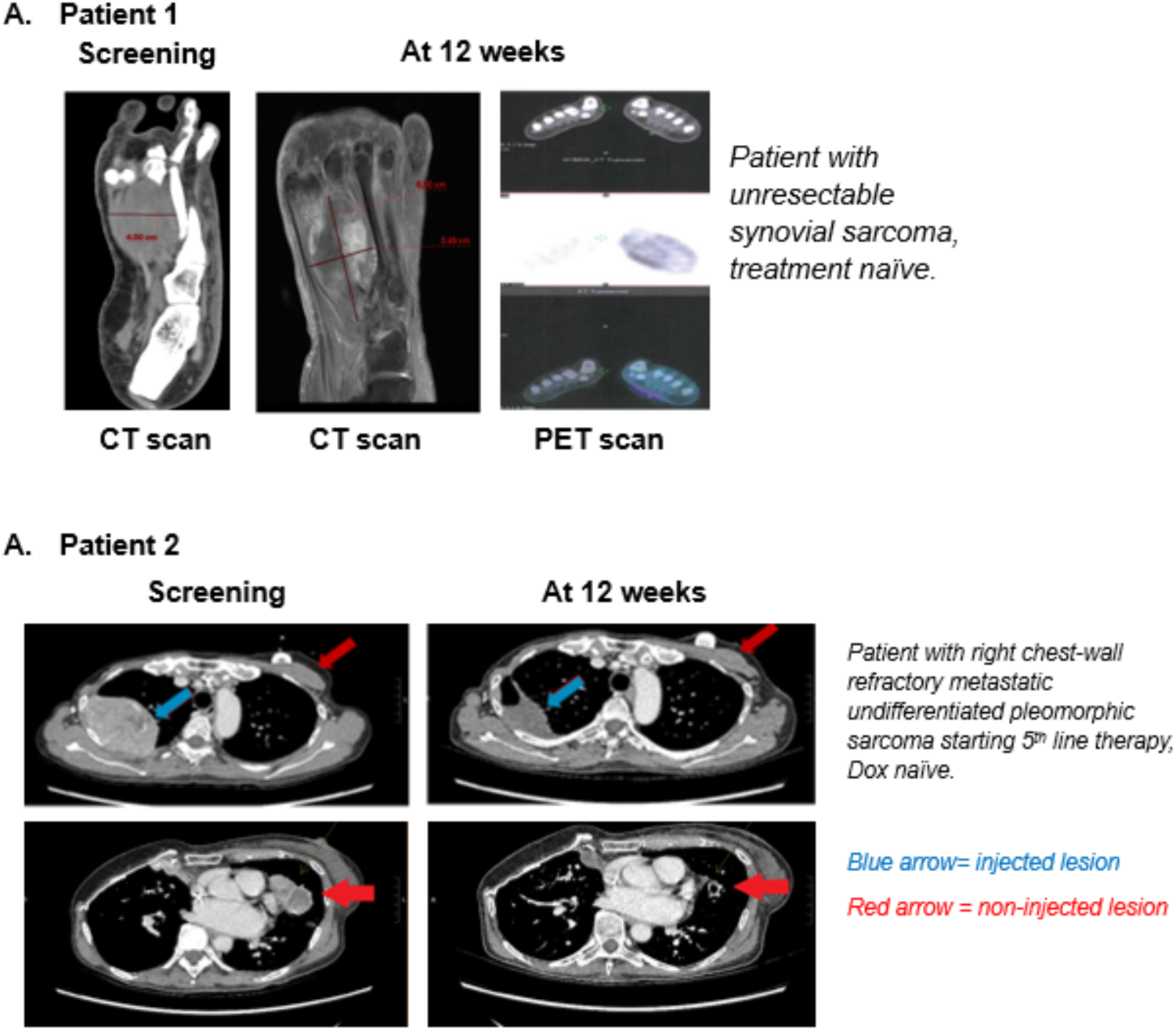
CT and PET scans for two Phase 2a cohort patients with unconfirmed partial response at screening (left image) and 12 Weeks (right image). CT, computed tomography; PET, positron emission tomography. Dox, doxorubicin.

### Systemic and tumor immune changes

Peripheral blood mononuclear cell (PBMC) samples from 39 patients (n=34 from Phase 1; n=5 from Phase 2a) were collected at baseline and before treatment cycles 2 and 3 and profiled by CyTOF. Similar results to the comprehensive analysis reported for the Phase 1 patient data^46^ were observed. This included a comparable composition of major circulating leukocyte populations between patients at baseline and after SQ3370 treatments (Fig 4A) as well as systemic changes that support a potential anti-tumor response related to antigen presentation and are reflective of a more immune-activated state, including an increase in circulating dendritic cells and central memory and terminal effector CD8^+^ T-cell subpopulations (Fig 4B).

**Fig 4.**
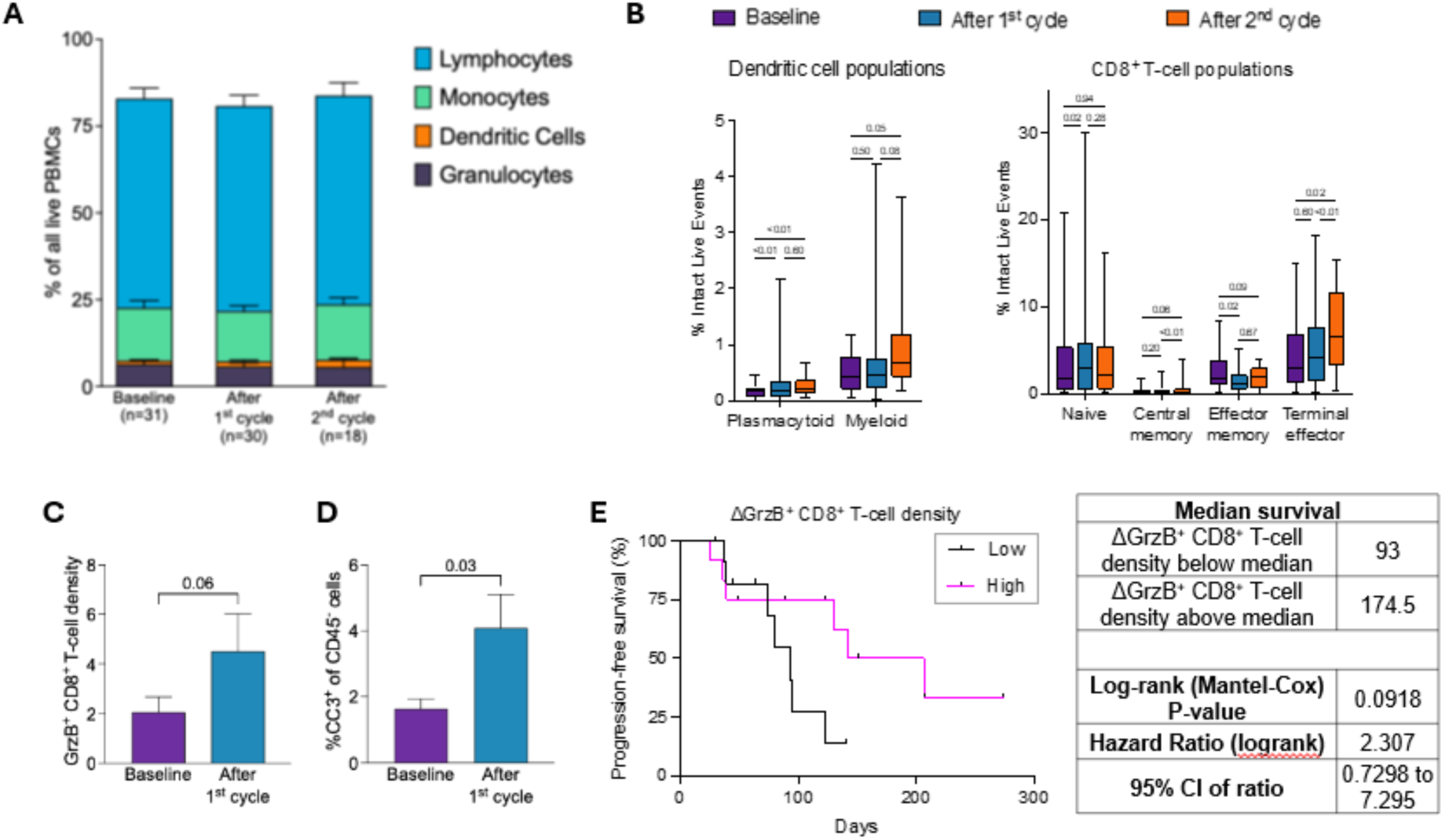
Immune profiling PBMCs and tumor biopsies from patient on SQ3370-001 clinical trial. **A-B.** PBMCs from 39 patients (34 Phase 1 and 5 Phase 2a; ≥2.8x Dox Eq) were profiled at baseline and after the initial two SQ3370 cycles using CyTOF analysis. A. Distribution of major immune cell types at different time points. No significance was observed between baseline and post-treatment time points. Stacked bar graph with mean ± SEM. *P*-value: Two-Way ANOVA, *post-hoc* Dunnett’s test vs baseline. B. Circulating CD8^+^ T-cell and dendritic cell subpopulations were assessed at three time points. Box plots with median, lower/upper quartiles; whiskers show minimum and maximum values. *P*-values: Mixed-effects model with Geisser-Greenhouse correction, *post-hoc* Tukey test. CD8^+^ T-cell subpopulations: naïve (CD8^+^CCR7^hi^CD45RA^+^CD45RO^-^), central memory (CD8^+^CCR7^hi^CD45RA^-^CD45RO^+^), effector memory (CD8^+^CCR7^lo/-^CD27^+^), terminal effector (CD8^+^CCR7^lo/-^CD27^-^). Dendritic cell subpopulations: plasmacytoid (HLA-DR^+^CD123^+^CD11c^-^); myeloid (HLA-DR^+^CD123^-^CD11c^+^CD38^+^). **C-D** Tumor microenvironment was assessed in biopsies from 33 patients (28 Phase 1, 5 Phase 2; ≥4x Dox Eq) at baseline and after one SQ3370 cycle using multiplex immunohistochemistry. C. Cytotoxic T-cell activity, quantified as cell density (cells/mm^2^) of Granzyme B-positive CD8+ T-cells (GrzB^+^CD45^+^CD3^+^CD8^+^). D. Cell death quantified as cleaved caspase 3-positive (CC3^+^) fraction of CD45^-^ cells (non-leukocytes). *P*-values: Mann-Whitney test. E. Progression-free survival of patients that were stratified by the median change in tumor GrzB^+^CD8^+^ T-cell densities (cells/mm^2^) after SQ3370 treatment vs baseline.

Tumor biopsies from 33 patients (n=28 from Phase 1; n=5 from Phase 2a) collected at baseline and before the second SQ3370 treatment cycle were analyzed by multiplex immunohistochemistry. Aligned with the PBMC analysis, changes in the tumor microenvironment observed in Phase 1 tumor samples^46^ were confirmed in the analysis that included the additional 5 Phase 2a patients. These changes included an increase in tumor cytotoxic activity as measured by Granzyme-B expression in CD8^+^ T-cells (GrzB^+^CD8^+^ T-cells) (Fig 4C) and an increase in tumor cell apoptosis as measured by cleaved caspase 3 expression among non-leukocytes (CC3^+^ CD45^-^ cells) (Fig 4D). Importantly, the extent of T-cell mediated cytotoxic activation correlated with improved clinical outcome, with patients who had a higher than median increase in GrzB^+^CD8^+^ T-cell density after SQ3370 experiencing longer PFS than patients whose change in GrzB^+^CD8^+^ T-cell density was lower than median (Fig 4E). These data highlight the importance of the systemic and tumor immune contexture in shaping SQ3370-mediated anti-tumor responses.

## DISCUSSION

We present clinical proof of concept for pre-targeting and selective drug activation at tumors using *in vivo* click chemistry. The CAPAC pre-targeting approach activates potent cancer therapies at tumors, increasing local active payload concentration while decreasing systemic toxicity; SQ3370 is the first investigational product to utilize *in vivo* click chemistry in humans.

SQ3370 treatment at the RP2D dose demonstrated activity across 14 patients with metastatic STS. There was a DCR of 71.4% (all ongoing at study termination). Objective clinical activity was observed in 3/14 (21.4%) patients, with one patient achieving stable disease and 2 patients achieving unconfirmed partial response at study termination. Interestingly, reductions in individual tumor sizes were observed in both injected and non-injected lesions, as had been seen in the preclinical models using SQ3370 ^37^. This, combined with the low rates of myleosuppression observed, suggests a shift from a myelosuppressive to a T-cell permissive tumor microenvironment, and systemic anti-tumor responses related to antigen presentation, reflective of a more immune-activated state, such as the expansion of dendritic cells and terminal effector and central memory CD8^+^ T-cells. Moreover, increased cytotoxic CD8^+^ T-cell activity and decreased myeloid-to-CD8^+^ T-cell ratio after SQ3370 treatment correlated with PFS, suggesting a connection between immunological effects of SQ3370 and improved clinical outcome, even among patients with STSs that are considered cold tumors. Overall, these data highlight the importance of the systemic and tumor immune contexture before and after SQ3370 treatment in shaping treatment-induced anti-tumor responses that may contribute to improved clinical outcomes. Further exploration is warranted to examine these enhanced immune effects.

The study was terminated at the recommendation of the sponsor and agreement of the Safety Review Committee. There were a number of reasons for this decision: 1) the study had achieved the objective of demonstrating proof of principle regarding the ability to use click chemistry in humans, the first time that this had been shown; 2) while the treatment was well tolerated from a safety perspective, the schedule of dosing represented a significant patient burden that impacted retention of patients on study. Despite this, most patients (54.7%; 29/53) received at least 4 treatment cycles of SQ3370; 15.5% (8/53) received at least 6 cycles, 5.7% (3/53) received at least 10 cycles, and 1.9% (1/53) received 12 cycles. Importantly, no differences were observed between those receiving 3 or 5 days of Dox protodrug, suggesting that a shorter dosing schedule could be explored with less burden on the patient and their families; 3) while 2 ongoing unconfirmed partial responses were observed in this predominantly metastatic population, the ORR was comparable to that observed with standard Dox, not meeting the prespecified criteria for study continuation. This suggests that dose escalation of Dox beyond a certain point is not associated with increased response rates.

Study limitations relate to the nature of an early Phase 1/2a study, enrolling a small cohort of heterogenous patients. Additionally, due to the early termination of the study, the long-term effects of SQ3370 in patients are still unknown. Doxorubicin is associated with significant cardiotoxicity. As such, clinicians limit the cumulative conventional Dox dose to 400–450 mg/m^2^; however, cardiac damage has been reported to occur at dosages considerably below this level. ^37,38,47,48^ In the current Phase 1/2a study, most patients (41/45 evaluable patients, 91.1%) received greater than conventional allowable cumulative dose of standard Dox (400 mg/m^2^) given as SQP33 protodrug. While cardiotoxicity was not dose-limiting in the current study, serious, irreversible heart problems may emerge. As such, evaluation of toxicity with high Dox concentrations remains of interest for SQ3370 therapy.

## CONCLUSION

Pre-targeting is an approach that prevents dose limiting toxicities associated with targeted therapies by limiting catabolic activation by normal tissues. The CAPAC pre-targeting approach activates potent cancer therapies at the tumor, increasing the payload concentration at the tumor while decreasing systemic toxicity. SQ3370 is the first reported use of *in vivo* click chemistry in humans and provides clinical validation of the CAPAC pre-targeting platform. The CAPAC platform is highly modular and can be applied to a variety of pre-targeting agents and payloads. Having demonstrated the ability to use click chemistry *in vivo* using an intratumorally injected pre-targeting agent in this study, we are now applying the technology to develop an ADC-like approach. This uses a clickable antigen binder (eg antibody or a smaller format) as a pre-targeting agent. Once bound at the tumor, this binder activates a clickable protodrug via the same click chemistry reaction employed by SQ3370 in this study. Importantly, both components can be administered systemically, minimizing patient burden. Our hypothesis is that this pre-targeting approach will reduce payload exposure and toxicity to normal cells, which has hampered the efficacy of ADCs to date.

## EHICAL APPROVAL

The name (site numbers) of institutional review boards (IRBs) that approved the protocol and all procedures are: Stanford University Research Compliance Office (1001), University of Texas: MD Anderson Cancer Center IRB (1002), WCG (1006, 1008, 1010, 1011), Oregon Health and Science University Institutional Review Board (1007), Bellberry Limited (5001, 5002, 5003).

## ACKNOWLEDGEMENTS

The authors thank all patients and their families for participating in SQ3370-001. The authors also thank the clinical teams for generating the data and for critical review of the manuscript (Sadie Whittaker, Ph.D., and Jesse McFarland, Ph.D.). Medical writing support was provided by Martha Mutomba, Ph.D., on behalf of Shasqi Inc.

## FUNDING

This work was supported by Shasqi, Inc and National Cancer Institute at the National Institutes of Health grants [1R44CA261573-01 and 1R44CA265665-01]. We select an open access license paid for by Shasqi, Inc.

## DISCLOSURE

S. P. Chawla has nothing to disclose. S. Abella, S. Wieland, T.-H. Nguyen, S. Srinivasan, M. Alečković, and J. M. Mejía Oneto are paid employees and shareholders of Shasqi Inc. S. Srinivasan holds stock or stock options in Tambo. J. M. Mejía Oneto is the Founder and CEO of Shasqi Inc, holds stock or stock options in Tambo, and has issued and pending patents for technology and compositions used in this study. V. Kwatra received honoraria from AstraZeneca, Bayer, and GSK; received support for attending meetings and/or travel from MSD; and received payment for participating on an advisory board for Jassen. V. Subbiah has nothing to disclose. N. Bui has nothing to disclose. V. Bhadri received support for attending meetings and/or travel from Shasqi Inc; received payment for participating on a medical advisory board for Dephera; and was a director for the Australia and New Zealand Sarcoma Association (unpaid). M. C. Weiss has nothing to disclose. M. Agulnik has nothing to disclose. C. W. Ryan has nothing to disclose. A. Guminski received research funding from Sun Pharma and AstraZeneca; received honoraria for an educational event from BMS and Sun Pharma; received support for attending meetings and/or travel from Sun Pharma and Astellas; and was on the advisory board for MSD.

## AUTHOR CONTRIBUTIONS

**S. P. Chawla:** Investigation, Resources, Writing - Review & Editing.

**E. Abella:** Conceptualization, Methodology, Data Curation, Writing - Original Draft, Writing - Review & Editing, Visualization.

**S. Wieland:** Conceptualization, Methodology, Formal analysis, Data Curation, Writing - Original Draft, Writing - Review & Editing, Visualization.

**T.-H. Nguyen:** Conceptualization, Methodology, Formal analysis, Data Curation, Writing - Original Draft, Writing - Review & Editing, Visualization.

**S. Srinivasan:** Conceptualization, Methodology, Formal analysis, Data Curation, Writing - Original Draft, Writing - Review & Editing, Visualization, Funding acquisition.

**M. Alečković:** Conceptualization, Methodology, Formal analysis, Data Curation, Writing - Original Draft, Writing - Review & Editing, Visualization.

**V. Kwatra:** Investigation, Resources, Writing - Review & Editing.

**V. Subbiah:** Investigation, Resources, Writing - Review & Editing.

**N. Bui:** Investigation, Resources, Writing - Review & Editing.

**V. Bhadri:** Investigation, Resources, Writing - Review & Editing.

**M. C. Weiss:** Investigation, Resources, Writing - Review & Editing.

**M. Agulnik:** Investigation, Resources, Writing - Review & Editing.

**C. W. Ryan:** Investigation, Resources, Writing - Review & Editing.

**A. D. Guminski:** Investigation, Resources, Writing - Review & Editing.

**J. M. Mejía Oneto:** Conceptualization, Methodology, Data Curation, Writing - Original Draft, Writing - Review & Editing, Visualization, Supervision, Project administration, Funding acquisition.

## TRIAL REGISTRATION

ClinicalTrials.gov registry, NCT04106492 (https://clinicaltrials.gov/ct2/show/NCT04106492).

## DATA AVAILABILITY

The data generated for this research are available within the article and supporting information. Data not shown will be made available upon request to the corresponding author.

## LIST OF SUPPLEMENTAL MATERIAL

Supplementary Figs S1 and S2

Supplementary Tables S1 to S7

## ABBREVIATIONS

ANOVA: Analysis of variance
AUC: Area under the curve
CAPAC: Click Activated Protodrugs Against Cancer
C_max_: Maximum concentration
DLT: Dose-limiting toxicity
Dox: Doxorubicin
Dox Eq: Doxorubicin molar equivalent
ECOG: Eastern Cooperative Oncology Group
H&E: Hematoxylin and eosin
IV: Intravenous
MALDI-MSI: Matrix-assisted laser desorption/ionization-imaging mass spectrometry
MTD: Maximum tolerated dose
NOAEL: No observed adverse effect level
PBMC: Peripheral blood mononuclear cells
QD: “quaque die”, once a day
RP2D: Recommended phase 2 dose
SQ3370: SQP33 protodrug + SQL70 biopolymer
SQL70: Intratumorally-injected biopolymer
SQP33: Doxorubicin protodrug
SC: Subcutaneous
TEAE: Treatment emergent adverse event

## Notes

### Competing Interest Statement

S. P. Chawla has nothing to disclose. S. Abella, S. Wieland, T.-H. Nguyen, S. Srinivasan, M. Alečković, and J. M. Mejia Oneto are paid employees and shareholders of Shasqi Inc. S. Srinivasan holds stock or stock options in Tambo. J. M. Mejia Oneto is the Founder and CEO of Shasqi Inc, holds stock or stock options in Tambo, and has issued and pending patents for technology and compositions used in this study.
V. Kwatra received honoraria from AstraZeneca, Bayer, and GSK; received support for attending meetings and/or travel from MSD; and received payment for participating on an advisory board for Jassen. V. Subbiah has nothing to disclose. N. Bui has nothing to disclose. V. Bhadri received support for attending meetings and/or travel from Shasqi Inc; received payment for participating on a medical advisory board for
Dephera; and was a director for the Australia and New Zealand Sarcoma Association (unpaid). M. C. Weiss has nothing to disclose. M. Agulnik has nothing to disclose. C. W. Ryan has nothing to disclose. A. Guminski received research funding from Sun Pharma and AstraZeneca; received honoraria for an educational event from BMS and Sun Pharma; received support for attending meetings and/or travel from Sun Pharma and Astellas; and was on the advisory board for MSD.

### Clinical Trial

NCT04106492

### Summary of Updates

Edited name of one author on the medRxiv submission portal. Previously shown as D. Scott Wieland. Now changed to Scott Wieland. No changes to the actual manuscript files.

## References

1 Jang SH, Wientjes MG, Lu D et al. Drug delivery and transport to solid tumors. Pharm Res 2003;20(9):1337–1350.

2 Minchinton AI, Tannock IF. Drug penetration in solid tumours. Nat Rev Cancer 2006;6(8):583–592.

3 Stuurman FE, Nuijen B, Beijnen JH et al. Oral anticancer drugs: mechanisms of low bioavailability and strategies for improvement. Clin Pharmacokinet 2013;52(6):399–414.

4 Livshits Z, Rao RB, Smith SW. An approach to chemotherapy-associated toxicity. Emerg Med Clin North Am 2014;32(1):167–203.

5 Juthani R, Punatar S, Mittra I. New light on chemotherapy toxicity and its prevention. BJC Reports 2024;2(1):41.

6 Basak D, Arrighi S, Darwiche Y et al. Comparison of Anticancer Drug Toxicities: Paradigm Shift in Adverse Effect Profile. Life (Basel) 2021;12(1).

7 Nurgali K, Jagoe RT, Abalo R. Editorial: Adverse Effects of Cancer Chemotherapy: Anything New to Improve Tolerance and Reduce Sequelae? Front Pharmacol 2018;9:245.

8 Bajjuri KM, Liu Y, Liu C et al. The legumain protease-activated auristatin prodrugs suppress tumor growth and metastasis without toxicity. ChemMedChem 2011;6(1):54–59.

9 Vaishampayan U, Glode M, Du W et al. Phase II study of dolastatin-10 in patients with hormone-refractory metastatic prostate adenocarcinoma. Clin Cancer Res 2000;6(11):4205–4208.

10 Buckel L, Savariar EN, Crisp JL et al. Tumor radiosensitization by monomethyl auristatin E: mechanism of action and targeted delivery. Cancer Res 2015;75(7):1376–1387.

11 Bai R, Pettit GR, Hamel E. Dolastatin 10, a powerful cytostatic peptide derived from a marine animal. Inhibition of tubulin polymerization mediated through the vinca alkaloid binding domain. Biochem Pharmacol 1990;39(12):1941–1949.

12 Nguyen TD, Bordeau BM, Balthasar JP. Mechanisms of ADC Toxicity and Strategies to Increase ADC Tolerability. Cancers (Basel) 2023;15(3).

13 Baah S, Laws M, Rahman KM. Antibody-drug conjugates-a tutorial review. Molecules 2021;26(10):2943.

14 Casi G, Neri D. Antibody-Drug Conjugates and Small Molecule-Drug Conjugates: Opportunities and Challenges for the Development of Selective Anticancer Cytotoxic Agents. J Med Chem 2015;58(22):8751–8761.

15 Baker MP, Reynolds HM, Lumicisi B et al. Immunogenicity of protein therapeutics: The key causes, consequences and challenges. Self Nonself 2010;1(4):314–322.

16 Herrera AF, Patel MR, Burke JM et al. Anti-CD79B Antibody-Drug Conjugate DCDS0780A in Patients with B-Cell Non-Hodgkin Lymphoma: Phase 1 Dose-Escalation Study. Clin Cancer Res 2022;28(7):1294–1301.

17 Palanca-Wessels MC, Czuczman M, Salles G et al. Safety and activity of the anti-CD79B antibody-drug conjugate polatuzumab vedotin in relapsed or refractory B-cell non-Hodgkin lymphoma and chronic lymphocytic leukaemia: a phase 1 study. Lancet Oncol 2015;16(6):704–715.

18 Pei Y, Li M, Hou Y et al. An autonomous tumor-targeted nanoprodrug for reactive oxygen species-activatable dual-cytochrome c/doxorubicin antitumor therapy. Nanoscale 2018;10(24):11418–11429.

19 Tap WD, Papai Z, Van Tine BA et al. Doxorubicin plus evofosfamide versus doxorubicin alone in locally advanced, unresectable or metastatic soft-tissue sarcoma (TH CR-406/SARC021): an international, multicentre, open-label, randomised phase 3 trial. Lancet Oncol 2017;18(8):1089–1103.

20 Gong J, Yan J, Forscher C et al. Aldoxorubicin: a tumor-targeted doxorubicin conjugate for relapsed or refractory soft tissue sarcomas. Drug Des Devel Ther 2018;12:777–786.

21 Mita MM, Natale RB, Wolin EM et al. Pharmacokinetic study of aldoxorubicin in patients with solid tumors. Invest New Drugs 2015;33(2):341–348.

22 Francisco JA, Cerveny CG, Meyer DL et al. cAC10-vcMMAE, an anti-CD30-monomethyl auristatin E conjugate with potent and selective antitumor activity. Blood 2003;102(4):1458–1465.

23 Liu Y, Bajjuri KM, Liu C et al. Targeting cell surface alpha(v)beta(3) integrin increases therapeutic efficacies of a legumain protease-activated auristatin prodrug. Mol Pharm 2012;9(1):168–175.

24 Li H, Yu C, Jiang J et al. An anti-HER2 antibody conjugated with monomethyl auristatin E is highly effective in HER2-positive human gastric cancer. Cancer Biol Ther 2016;17(4):346–354.

25 DeFeo-Jones D, Brady SF, Feng DM et al. A prostate-specific antigen (PSA)-activated vinblastine prodrug selectively kills PSA-secreting cells in vivo. Mol Cancer Ther 2002;1(7):451–459.

26 DeFeo-Jones D, Garsky VM, Wong BK et al. A peptide-doxorubicin ‘prodrug’ activated by prostate-specific antigen selectively kills prostate tumor cells positive for prostate-specific antigen in vivo. Nat Med 2000;6(11):1248–1252.

27 DiPaola RS, Rinehart J, Nemunaitis J et al. Characterization of a novel prostate-specific antigen-activated peptide-doxorubicin conjugate in patients with prostate cancer. J Clin Oncol 2002;20(7):1874–1879.

28 Albright CF, Graciani N, Han W et al. Matrix metalloproteinase-activated doxorubicin prodrugs inhibit HT1080 xenograft growth better than doxorubicin with less toxicity. Mol Cancer Ther 2005;4(5):751–760.

29 Verhoeven M, Seimbille Y, Dalm SU. Therapeutic Applications of Pretargeting. Pharmaceutics 2019;11(9).

30 Poty S, Ordas L, Dekempeneer Y et al. Optimizing the Therapeutic Index of sdAb-Based Radiopharmaceuticals Using Pretargeting. J Nucl Med 2024;65(10):1564–1570.

31 The Royal Swedish Academy of Sciences. Click chemistry and bioorthogonal chemisty. The Noble Prize Committee of Chemistry. 5 October 2022. https://www.nobelprize.org/uploads/2022/10/advanced-chemistryprize2022-2.pdf. Accessed 11 March 2025.

32 Scinto SL, Bilodeau DA, Hincapie R et al. Bioorthogonal chemistry. Nat Rev Methods Primers 2021;1.

33 Blackman ML, Royzen M, Fox JM. Tetrazine ligation: fast bioconjugation based on inverse-electron-demand Diels-Alder reactivity. J Am Chem Soc 2008;130(41):13518–13519.

34 Devaraj NK, Thurber GM, Keliher EJ et al. Reactive polymer enables efficient in vivo bioorthogonal chemistry. Proc Natl Acad Sci U S A 2012;109(13):4762–4767.

35 Jewett JC, Bertozzi CR. Cu-free click cycloaddition reactions in chemical biology. Chem Soc Rev 2010;39(4):1272–1279.

36 McFarland JM, Alečković M, Coricor G et al. Click chemistry selectively activates an auristatin protodrug with either intratumoral or systemic tumor-targeting agents. ACS Cent Sci 2023;9(7):1400–1408.

37 Srinivasan S, Yee NA, Wu K, et al. SQ3370 activates cytotoxic drug via click chemistry at tumor and elicits sustained responses in injected & non-injected lesions. Adv Ther (Weinh) 2021;4(3):2000243.

38 Wu K, Yee NA, Srinivasan S et al. Click activated protodrugs against cancer increase the therapeutic potential of chemotherapy through local capture and activation. Chem Sci 2021;12(4):1259–1271.

39 von Mehren M, Kane JM, Agulnik M, et al. Soft Tissue Sarcoma, Version 2.2022, NCCN Clinical Practice Guidelines in Oncology. J Natl Compr Canc Netw 2022;20(7):815–833.

40 Lahat G, Tuvin D, Wei C et al. New perspectives for staging and prognosis in soft tissue sarcoma. Ann Surg Oncol 2008;15(10):2739–2748.

41 Sbaraglia M, Dei Tos AP. The pathology of soft tissue sarcomas. Radiol Med 2019;124(4):266–281.

42 National Comprehensive Cancer Network. National Comprehensive Cancer Network, NCCN Clinical Practice Guidelines in Oncology (NCCN Guidelines®). Soft Tissue Sarcoma. Version 5.2024 — March 10, 2025. https://www.nccn.org/professionals/physician_gls/pdf/sarcoma.pdf. Accessed 11 March 2025.

43 Seddon B, Strauss SJ, Whelan J et al. Gemcitabine and docetaxel versus doxorubicin as first-line treatment in previously untreated advanced unresectable or metastatic soft-tissue sarcomas (GeDDiS): a randomised controlled phase 3 trial. Lancet Oncol 2017;18(10):1397–1410.

44 Judson I, Verweij J, Gelderblom H et al. Doxorubicin alone versus intensified doxorubicin plus ifosfamide for first-line treatment of advanced or metastatic soft-tissue sarcoma: a randomised controlled phase 3 trial. Lancet Oncol 2014;15(4):415–423.

45 Ryan CW, Merimsky O, Agulnik M et al. PICASSO III: A Phase III, Placebo-Controlled Study of Doxorubicin With or Without Palifosfamide in Patients With Metastatic Soft Tissue Sarcoma. J Clin Oncol 2016;34(32):3898–3905.

46 Srinivasan S, A. Yee N, Zakharian M et al. SQ3370, the first clinical click chemistry-activated cancer therapeutic, shows safety in humans and translatability across species. 2023:bioRxiv [Preprint]. 2023 Mar 2029:2023.2003.2028.534654. doi: 534610.531101/532023.534603.534628.534654. https://www.biorxiv.org/content/534610.531101/532023.534603.534628.534654v534651.full.pdf. Accessed 25 April 2025. In press at Clin Cancer Res.

47 Barrett-Lee PJ, Dixon JM, Farrell C et al. Expert opinion on the use of anthracyclines in patients with advanced breast cancer at cardiac risk. Ann Oncol 2009;20(5):816–827.

48 Vitfell-Rasmussen J, Krarup-Hansen A, Vaage-Nilsen M et al. Real-life incidence of cardiotoxicity and associated risk factors in sarcoma patients receiving doxorubicin. Acta Oncol 2022;61(7):801–808.

49 Tsujikawa T, Kumar S, Borkar RN et al. Quantitative multiplex immunohistochemistry reveals myeloid-inflamed tumor-immune complexity associated with poor prognosis. Cell Rep 2017;19(1):203–217.

