## Supplement for "A first-in-human, Phase 1/2a, open-label study of SQ3370, a first-in-class doxorubicin-based click chemistry therapeutic, in patients with advanced solid tumors"

**Title**

**Author Institutions**

[1]Sarcoma Oncology Research Center, Santa Monica, CA, USA

[2]Shasqi Inc, San Francisco, CA, USA

[3]Cancer Research South Australia, Adelaide, Australia

[4]Department of Investigational Cancer Therapeutics, Division of Cancer Medicine, The University of Texas MD Anderson Cancer Center, Houston, TX, USA

[5]Stanford University, Stanford, CA, USA

[6]Chris O'Brien Lifehouse, Sydney, Australia

[7]Washington University in St Louis, St Louis, MO, USA

[8]Department of Medical Oncology & Therapeutics Research, City of Hope, Duarte, CA, USA

[9]Knight Cancer Institute, Oregon Health & Science University, Portland, OR, USA

[10]Department of Medical Oncology, Royal North Shore Hospital, St Leonards, Australia

### Supplementary Methods

#### *Sample size*

Sample size for the Phase 1 dose escalation portion of the study was not predefined; total enrollment depended on the DLTs observed and number of escalation cohorts. For the Extremity STS Group 1, sample size was guided by Simon 2-stage (optimal) designs based on historical objective responses per group. The Unresectable STS Group 2 utilized the Continuous Reassessment Method design. Different criteria were applied to determine the number of participants for each stage and the strength of the efficacy signal that would lead to a recommendation of proceeding to the next stage. In the Extremity STS Group 1, up to approximately 28 patients were to be treated in Stage 1. In the Unresectable STS Group 2, a total of 22 patients were to be treated with a planned interim after the first 12 patients were enrolled. For all Phase 1 dose escalation cohorts, patients could be replaced if they were enrolled into the study but did not receive the intended dose of SQL70 biopolymer or SQP33 protodrug during cycle 1 (for any reason other than TEAEs or serious TEAEs). If cycle 1 could not be completed due to a COVID-19 infection, an additional patient could be enrolled in the group.

#### *SQ3370 administration*

SQL70 biopolymer was administered intra and/or peritumorally on day 1 of each 21-day cycle into a lesion or lesions at a fixed 10 mL or 20 mL injection volume, followed by 3 or 5 consecutive daily infusions of SQP33 protodrug (day 1 to day 3 or day 5) with the cycle repeated 21 days after initiation of the previous cycle (Supplementary Fig S1). SQP33 protodrug was administered as described below.

##### *Phase 1 dose escalation*

In the Phase 1 dose escalation portion of the study, patients received specific dose levels of SQP33 protodrug, assigned according to the dose escalation design (Supplementary Fig S1), which comprises of an initial period of accelerated titration cohorts (Stage 1) followed by 3+3 (Rolling 6) cohorts (Stage 2) (Simon 1997). For the Stage 1 accelerated titration step, 1 patient was enrolled per cohort with a doubling of the SQP33 protodrug dose level between cohorts. For the Stage 2 3+3 (rolling 6) step, 3-6 patients were concurrently enrolled per cohort to determine the dose level increase and minimum number of patients enrolled per dose level. Dose-level assignments were based on the number of patients currently enrolled in the cohort, the number of DLTs observed, the number of patients who had not completed the DLT period, general tolerability observed, and any available pharmacokinetic data.

Patients received the SQL70 biopolymer in 3 separate groups. In the 10 mL SQL70 biopolymer cohorts 1-9, SQL70 biopolymer was administered intra and/or peritumorally at a fixed 10 mL injection volume. The starting daily dose of SQP33 protodrug for this clinical study was 8 mg/m$^2$/day (5.72 mg/m$^2$/day Dox equivalent [Eq]). If the injected lesion was considered no longer injectable by the investigator, then the area of the tumor bed, if possible, was treated for 2 additional cycles. Additional lesion(s) could be treated with a further 10 mL of SQL70 biopolymer (for a total of 20 mL SQL70 biopolymer maximum administered per cycle) as considered appropriate by the investigator. In the 20 mL SQL70 biopolymer (from Cycle 1) cohort to evaluate the administration of 20 mL of SQL70 biopolymer in 1 or 2 lesions starting at Cycle 1, patients were enrolled into a separate cohort at a dose level of SQP33 protodrug that was lower than the current dose level being evaluated. In the 20 mL SQL70 biopolymer (post-Cycle 1) cohort, patients on treatment who attained at least a 30% reduction in injected tumor size or had a tumor that is no longer injectable could have a second lesion injected with an additional 10 mL of SQL70 biopolymer, thereby receiving a total of 20 mL of SQL70 biopolymer.

The SRC reviewed the available safety data at the completion of Cycle 1 for each of the dose escalation cohorts and the cohort receiving 20 mL SQL70 biopolymer from Cycle 1 to evaluate possible DLTs, recommend whether more patients should be enrolled at a given dose level or cohort, and recommend whether to dose escalate or to stop enrollment based on the definition of DLTs in addition to its members' best clinical judgement.

*Phase 2 expansion groups*

The Phase 2a consisted of two dose escalation treatment groups of Extremity STS Group 1 and the Unresectable STS Group 2 (Supplementary Fig S1), with patients receiving up to 12 cycles. No patients were enrolled in the originally planned Head and Neck Group 3.

The Extremity STS Group 1 cohort included treatment naïve patients with STS of the extremity American Joint Committee on Cancer (AJCC) Stage III or IV (> 5 cm injectable tumors) locally advanced and or metastatic tumors, ineligible for primary surgical intervention, according to the consensus of a multidisciplinary treatment team, determined prior to screening (Supplementary Fig S1). Procedure for Extremity STS Group 1 was conducted in 2 parts with a possible expansion part. The group opened with the initial phase safety run-in of 6 patients to establish the safety and initial feasibility of a 12-week delay in definitive surgical resection in patients with high-risk STS of the extremity treated with SQ3370, and 31 additional patients initial adaptive design phase. At completion of the 6 patient safety run-in, the Extremity STS Group 1 cohort followed a Simon 2-stage design.

The Unresectable STS Group 2 cohort included Dox naïve patients with unresectable STS locally advanced or metastatic cancers with prior first- or second-line therapy (Supplementary Fig S1). Patients were enrolled at the RP2D of SQ3370, maintaining the total dose per cycle constant to compare 3-day vs standard 5-day protodrug infusion schedules. The 3-day SQP33 protodrug infusion schedule administered the same protodrug dose on day 1 as the 5-day protodrug infusion schedule and 2x the doses on days 2 and 3 of the 5-day schedule, with up to a total of 11 patients per group planned. Patients were treated with the RP2D of SQ3370 every 21 days (1 cycle) up to 12 cycles or until other treatment discontinuation criteria were met. On day 1 of each cycle, the patient received an injection of SQL70 biopolymer into the tumor (10 mL) followed by 3 or 5 days of SQP33 infusion starting 3 hours ± 30 minutes after the SQL70 biopolymer injection.

#### ***Study assessments***

*Pharmacokinetics*

Plasma samples for determination of pharmacokinetic levels of SQP33 protodrug, SQL70 biopolymer and active Dox following SQ3370 treatment were withdrawn pre-infusion of SQ3370 and then at specified times post-infusion SQ3370on days 1-5 of all cycles. Plasma pharmacokinetic bioanalysis was conducted using validated liquid chromatography-mass spectrometry method to determine maximum plasma concentration ($C_{max}$), timepoint of maximum observed plasma concentration ($T_{max}$), and area under the plasma concentration curve from time zero to the last measurable time point for individual days and/or complete cycles ($AUC_t$).

*Tumor assessments*

Tumor response was evaluated using RECIST 1.1 based on contrast-enhanced computed tomography (CT) or magnetic resonance imaging (MRI).

*Immune cell profiling of patient blood and tumor samples*

Immune analysis of PBMCs and tumor biopsies of patients at baseline and after the initial two SQ3370 cycles were conducted as previously published.[46] PBMCs were successfully collected and analyzed from 39 patients (34 from the Phase 1 arm and 5 from the Phase 2a arm; all treated with ≥2.8x Dox Eq) using CyTOF analysis. This included a total of 31 baseline, 30 second, and 18 third time point samples. Only samples with more than 100,000 viable cells were included in the analysis. Briefly, PBMCs were stained with the MaxPar Direct Immune Profiling Assay kit (Standard Biotools) and run on the Helios mass cytometer (Standard Biotools), with the resulting normalized .fcs files being analyzed using the Maxpar Pathsetter software (Standard Biotools).[46]

The tumor microenvironment was assessed in tumor biopsies from 33 patients (28 from the Phase 1 arm, 5 from the Phase 2 arm; all treated with ≥4x Dox Eq) at baseline and after one SQ3370 cycle using multiplex immunohistochemistry. Briefly, sequential immunohistochemistry was performed on 5 μm formalin-fixed paraffin embedded (FFPE) sections using an adapted protocol based on methodology previously described,[49] using the following antibody staining conditions: primary antibody incubations were carried out for 1 hour at 4 °C using CC3 (ASP175, 1:400, Cell Signaling; RRID:AB_2341188); and 30 min at room temperature using CD45 (H130, 1:100, eBioscience; RRID:AB_467274), CD3 (SP7, 1:150, Thermo Fisher Scientific; RRID: AB_10982026), CD8 (SP16, 1:100, Thermo Fisher Scientific; RRID: AB_10984334), or GRZB (Polyclonal, 1:200, Abcam; RRID: AB_304251). For secondary antibody incubations, either anti-rat, anti-mouse, or anti-rabbit Histofine Simple Stain MAX PO horseradish peroxidase (HRP)-conjugated polymer (Nichirei Biosciences, Tokyo, Japan) was applied for 30 min at room temperature, followed by AEC chromogen (Vector Laboratories, Burlingame, CA, USA). Slides were digitally scanned following each chromogen development, and the staining process was repeated for all subsequent staining cycles. Multiplex IHC image processing was performed as previously reported (DOI: dx.doi.org/10.17504/protocols.io.n92ldmmznl5b/v2).

**Supplementary Fig S1:** Treatment schematic, single patient accelerated dose escalation followed by 3+3 design with modified Fibonacci dose escalation in the Phase 1 portion of the study, and dose expansion in Extremity STS Group 1 cohort and unresectable STS Group 2 cohort and Group 3 head and neck cohort in the Phase 2a portion of the study.

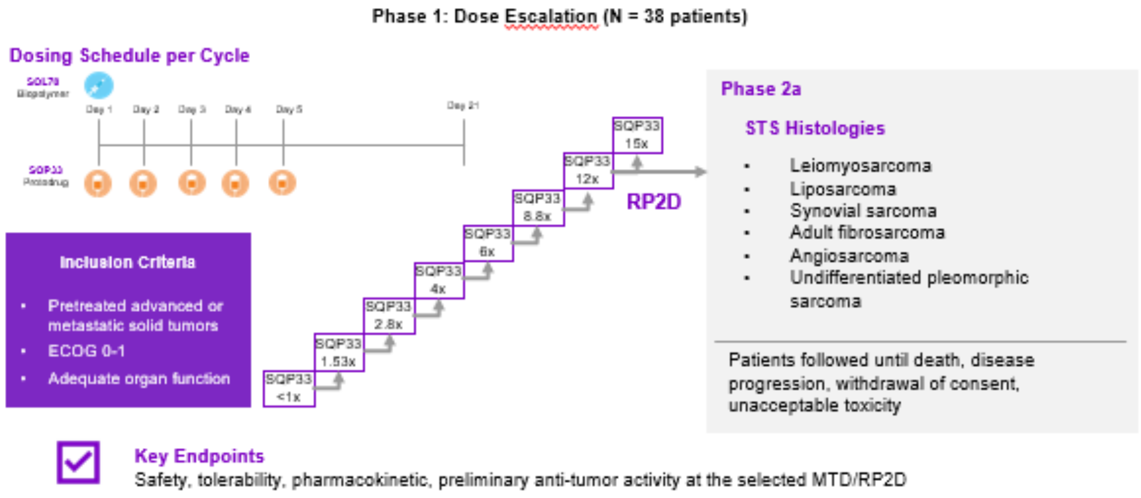


**Phase 2a**

| Group | Treatment Line | Disease State | Design | Purpose | Population | Treatment Plan |
|---|---|---|---|---|---|---|
| 1 | 1st | Locally advanced and/or metastatic extremity sarcoma (STS) | Simon 2 Stage with safety run-in | Define activity in earlier line of treatment | Treatment-naïve ineligible for primary surgery (1 line) | 4-cycle induction with 5-day schedule |
| 2 | 1st or 2nd | Unresectable and/or metastatic STS | Continuous reassessment | Optimize schedule for further development | Dox-naïve locally advance +/- prior Rx (1 or 2nd line) | Compare 3-day vs 5-day schedule |

One treatment cycle of SQ3370 consisted of a 10 mL or 20 mL SQL70 biopolymer injection on day 1, followed by SQP33 protodrug infusion on day 1 through day 3 or day 5 (3 or 5 doses total). The cycle was repeated 21 days after initiation of the previous cycle. SQP33 dose #x = # times doxorubicin molar equivalence. AE, adverse event; DLT, dose limiting toxicity; ECOG, Eastern Cooperative Oncology Group; MTD, maximum tolerated dose; RP2D, recommended phase 2 dose; STS, soft tissue sarcoma.

One treatment cycle of SQ3370 consisted of a 10 mL or 20 mL SQL70 biopolymer injection on day 1, followed by SQP33 protodrug infusion on day 1 through day 3 or day 5 (3 or 5 doses total). The cycle was repeated 21 days after initiation of the previous cycle. SQP33 dose #x = # times doxorubicin molar equivalence. AE, adverse event; DLT, dose limiting toxicity; ECOG, Eastern Cooperative Oncology Group; MTD, maximum tolerated dose; RP2D, recommended phase 2 dose; STS, soft tissue sarcoma.

**Supplementary Fig S2.** Patient disposition[a]

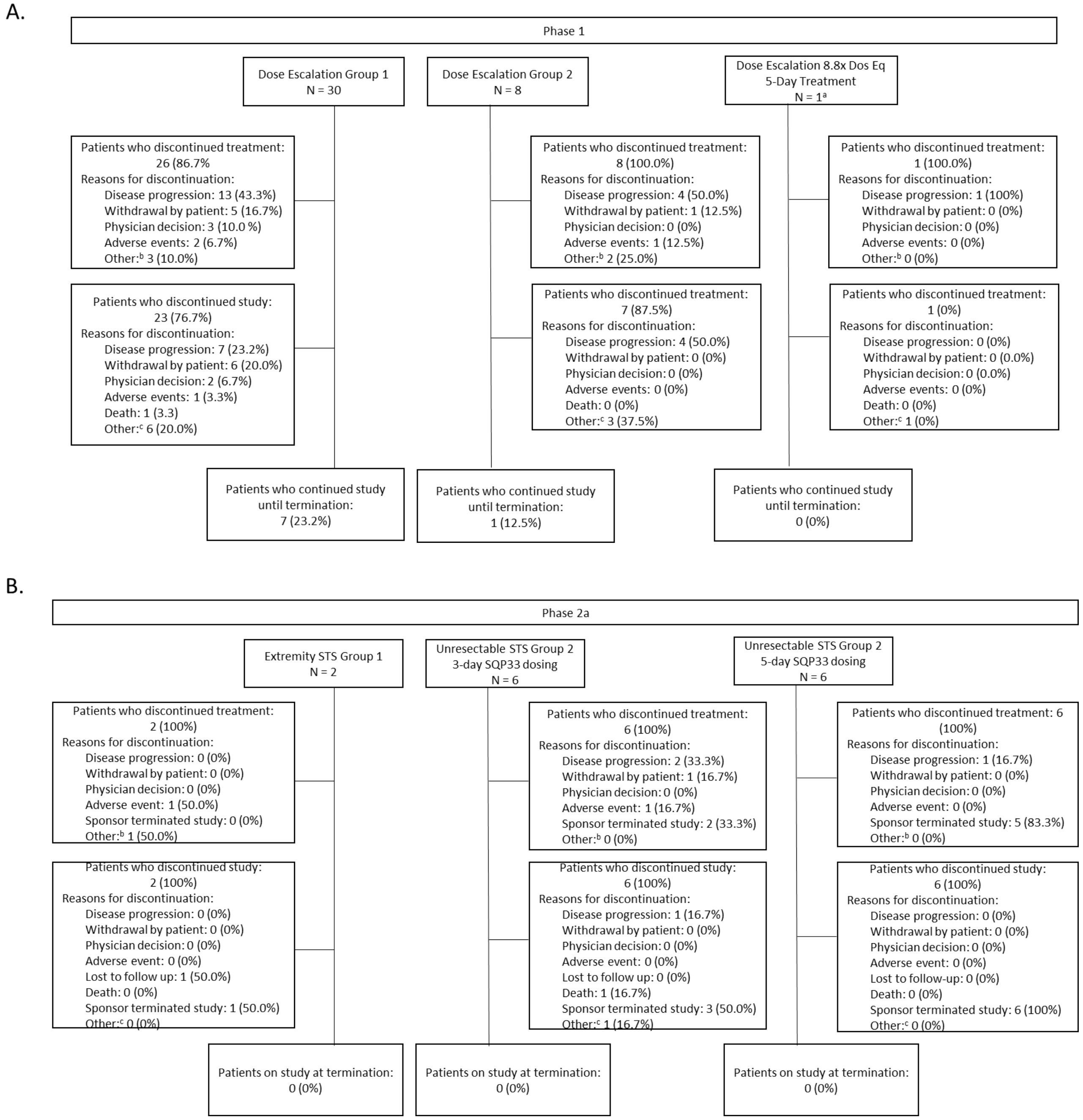


Data cutoff: 15 December 2022 for the Phase 1 cohorts and 07 September 2023 for the Phase 2 cohorts.
N = number of patients who received ≥1 dose of SQL70 biopolymer and 1 dose of SQP33 protodrug.
[a]1 patient who received 8.8x standard Dox dose was not reported as part of the Phase 1 cohorts.
[b]Reasons for treatment discontinuation in 3 patients in the Dose Escalation Group 1 were disease progression (2 patients) and disease-related death (1 patient); in 2 patients in the Dose Escalation Group 2 were pregnancy (1 patient) and disease progression (1 patient); and in 1 patient in the Extremity STS Group 1 was surgical resection of tumor that led to treatment discontinuation.
[c]Reasons for study discontinuation in 6 patients in the Dose Escalation Group 1 were disease progression (2 patients) and death due to disease progression (3 patients), and death (1 patient); in 3 patients in the Dose Escalation Group 2 were death due to disease progression (1 patient), pregnancy (1 patient) and transferred to

hospice care were the patient died (1 patient); in 1 patient in the 8.8x Dox Eq dose was planned for another treatment regimen; and in 1 patient in the Unresectable STS Group 2 3-day dosing schedule was trial closure. Dox Eq, doxorubicin molar equivalents.

**Supplementary Table S1:** Patient disposition, treatment and study duration in the Phase 1 Dose Escalation Group 1 cohort (Safety Population)

| | **10 mL SQL70 Biopolymer + SQP33 Dox Eq Dose (mg/m²)** | | | | | | | | | |
|---|---|---|---|---|---|---|---|---|---|---|
| | **0.38x (N = 1)** | **0.76x (N = 1)** | **1.53x (N = 1)** | **2.8x (N = 3)** | **4x (N = 3)** | **6x (N = 4)** | **8.8x (N = 4)** | **12x (N = 7)** | **15x (N = 6)** | **Total (N = 30)** |
| Number of treatment discontinuations, n (%) | 1 (100.0) | 1 (100.0) | 1 (100.0) | 3 (100.0%) | 3 (100.0) | 4 (100.0) | 4 (100.0) | 7 (100.0) | 2 (33.3) | 26 (86.7) |
| Reason for treatment discontinuation, n (%) | | | | | | | | | | |
| Disease progression | 0 (0.0) | 1 (100.0) | 0 (0.0) | 2 (66.7) | 1 (33.3) | 2 (50.0) | 3 (75.0) | 2 (28.6) | 2 (33.3) | 13 (43.3) |
| Adverse event | 0 (0.0) | 0 (0.0) | 0 (0.0) | 0 (0.0) | 0 (0.0) | 0 (0.0) | 0 (0.0) | 2 (28.6) | 0 (0.0) | 2 (6.7) |
| Physician decision | 0 (0.0) | 0 (0.0) | 0 (0.0) | 1 (33.3) | 0 (0.0) | 1 (25.0) | 0 (0.0) | 1 (14.3) | 0 (0.0) | 3 (10.0) |
| Withdrawal by subject | 0 (0.0) | 0 (0.0) | 1 (100.0) | 0 (0.0) | 1 (33.3) | 1 (25.0) | 1 (25.0) | 1 (14.3) | 0 (0.0) | 5 (16.7) |
| Other | 1 (100.0) | 0 (0.0) | 0 (0.0) | 0 (0.0) | 1 (33.3) | 0 (0.0) | 0 (0.0) | 1 (14.3) | 0 (0.0) | 3 (10.0) |
| Number of study discontinuations, n (%) | 1 (100.0) | 1 (100.0) | 1 (100.0) | 3 (100.0%) | 3 (100.0) | 3 (75.0%) | 3 (75.0) | 7 (100.0) | 1 (16.7) | 23 (76.7) |
| Reason for study discontinuation, n (%) | | | | | | | | | | |
| Disease progression | 0 (0.0) | 1 (100.0) | 0 (0.0) | 2 (66.7) | 1 (33.3) | 0 (0.0) | 1 (25.0) | 1 (14.3) | 1 (16.7) | 7 (23.3) |
| Adverse event | 0 (0.0) | 0 (0.0) | 0 (0.0) | 0 (0.0) | 0 (0.0) | 0 (0.0) | 0 (0.0) | 1 (14.3) | 0 (0.0) | 1 (3.3) |
| Physician decision | 0 (0.0) | 0 (0.0) | 0 (0.0) | 1 (33.3) | 0 (0.0) | 0 (0.0) | 0 (0.0) | 1 (14.3) | 0 (0.0) | 2 (6.7) |
| Withdrawal by subject | 0 (0.0) | 0 (0.0) | 1 (100.0) | 0 (0.0) | 1 (33.3) | 1 (25.0) | 1 (25.0) | 2 (28.6) | 0 (0.0) | 6 (20.0) |
| death | 1 (100.0) | 0 (0.0) | 0 (0.0) | 0 (0.0) | 0 (0.0) | 0 (0.0) | 0 (0.0) | 0 (0.0) | 0 (0.0) | 1 (3.3) |
| Other | 0 (0.0) | 0 (0.0) | 0 (0.0) | 0 (0.0) | 1 (33.3) | 2 (50.0) | 1 (25.0) | 2 (28.6) | 0 (0.0) | 6 (20.0) |

| | 10 mL SQL70 Biopolymer + SQP33 Dox Eq Dose (mg/m$^2$) | | | | | | | | | |
|---|---|---|---|---|---|---|---|---|---|---|
| | **0.38x (N = 1)** | **0.76x (N = 1)** | **1.53x (N = 1)** | **2.8x (N = 3)** | **4x (N = 3)** | **6x (N = 4)** | **8.8x (N = 4)** | **12x (N = 7)** | **15x (N = 6)** | **Total (N = 30)** |
| Number ongoing, n (%) | 0 (0.0) | 0 (0.0) | 0 (0.0) | 0 (0.0) | 0 (0.0) | 1 (25.0) | 1 (25.0) | 0 (0.0) | 5 (83.3) | 7 (23.3) |
| Time on study (days) | | | | | | | | | | |
| Mean (SD) | 148 | 51 | 85 | 53 ± 25 | 196 ± 67 | 159 ± 162 | 199 ± 147 | 70 ± 46 | 58 ± 10 | 109 ± 97 |
| Median, (Min, Max) | 148 (148, 148) | 51 (51, 51) | 85 (85, 85) | 65 (25, 70) | 207 (124, 257) | 101.5 (40, 393) | 192 (54, 333) | 64 (16, 149) | 58 (40, 69) | 68 (16, 393) |

Data cutoff: 15 December 2022.

N = number of patients who received ≥1 dose of SQL70 biopolymer and 1 dose of SQP33 protodrug.

Dox Eq, doxorubicin molar equivalents; Max, maximum; Min, minimum; SD, standard deviation.

**Supplementary Table S2:** Patient disposition, treatment and study discontinuation in the Phase 1 Dose Escalation Group 2 cohort (Safety Population)

| | **20 mL SQL70 Biopolymer + SQP33 Dox Eq Dose (mg/m²)** | | |
|---|---|---|---|
| | **4x (N=5)** | **6x (N=3)** | **Total (N=8)** |
| Number of treatment discontinuations, n (%) | 5 (100.0%) | 3 (100.0%) | 8 (100.0%) |
| Reason for treatment discontinuation, n (%) | | | |
| Disease progression | 3 (60.0%) | 1 (33.3%) | 4 (50.0%) |
| Adverse Event | 1 (20.0%) | 0 (0.0%) | 1 (12.5%) |
| Physician decision | 0 (0.0%) | 0 (0.0%) | 0 (0.0%) |
| Withdrawal by subject | 0 (0.0%) | 1 (33.3%) | 1 (12.5%) |
| Other | 1 (20.0%) | 1 (33.3%) | 2 (25.0%) |
| Number of study discontinuations, n (%) | 5 (100.0%) | 2 (66.7%) | 7 (87.5%) |
| Reason for study discontinuation, n (%) | | | |
| Disease progression | 3 (60.0%) | 1 (33.3%) | 4 (50.0%) |
| Adverse event | 0 (0.0%) | 0 (0.0%) | 0 (0.0%) |
| Physician decision | 0 (0.0%) | 0 (0.0%) | 0 (0.0%) |
| Withdrawal by subject | 0 (0.0%) | 0 (0.0%) | 0 (0.0%) |
| Death | 0 (0.0%) | 0 (0.0%) | 0 (0.0%) |
| Other | 2 (40.0%) | 1 (33.3%) | 3 (37.5%) |
| Number ongoing, n (%) | 0 (0.0%) | 1 (33.3%) | 1 (12.5%) |
| Time on study (days) | | | |
| Mean (SD) | 80.8 ± 30.06 | 70.7 ± 63.71 | 77 ± 41.27 |
| Median (Min, Max) | 71 (45, 114) | 39 (29, 144) | 67.5 (29, 144) |

Data cutoff: 15 December 2022.
N = number of patients who received ≥1 dose of SQL70 biopolymer and 1 dose of SQP33 protodrug.
Dox Eq, doxorubicin molar equivalents; Max, maximum; Min, minimum; SD, standard deviation.

### Supplementary Table S3: SQ3370 administration and exposure for the Phase 1 Dose Escalation Group 1 (safety population)

| | 10 mL SQL70 Biopolymer + SQP33 Dox Eq Dose (mg/m$^2$) | | | | | | | | | |
|---|---|---|---|---|---|---|---|---|---|---|
| | **0.38x (N=1)** | **0.76x (N=1)** | **1.53x (N=1)** | **2.8x (N=3)** | **4x (N=3)** | **6x (N=4)** | **8.8x (N=4)** | **12x (N=7)** | **15x (N=6)** | **Total (N=30)** |
| Duration of SPQ3370 therapy (days) | | | | | | | | | | |
| Mean | 110 | 26 | 66 | 26.3 | 173 | 131 | 100.3 | 40.1 | 47.2 | 76.3 |
| SD | 18.9 | 84 | 159.01 | 83.6 | 35.78 | 17.39 | 80.66 | SD | 18.9 | 84 |
| Median | 110 | 26 | 66 | 33 | 173 | 68 | 83.5 | 33 | 54 | 49 |
| (Min, Max) | (110, 110) | (26, 26) | (66, 66) | (5, 41) | (89, 257) | (26, 362) | (26, 208) | (2, 103) | (26, 69) | (2, 362) |
| Number of SQL70 cycles started | | | | | | | | | | |
| 1 | | | | 1 (33.3%) | | | | 2 (28.6%) | | 3 (10.0%) |
| 2 | | 1 (100.0%) | | 2 (66.7) | | 2 (50.0) | 1 (25.0%) | 3 (42.9%) | 2 (33.3%) | 11 (36.7%) |
| 3 | | | | | | | 1 (25.0) | | 3 (50.0) | 4 (13.3) |
| 4 | | | 1 (100.0) | | | | | 2 (28.6%) | 1 (16.7%) | 4 (13.3%) |
| 5 | | | | | 1 (33.3) | | | | | 1 (3.3) |
| 6 | 1 (100.0) | | | | | 1 (25.0) | 1 (25.0) | | | 3 (10.0) |
| 9 | | | | | 1 (33.3) | | | | | 3 (10.0) |
| 10 | | | | | | | 1 (25.0) | | | 1 (3.3) |
| 12 | | | | | 1 (33.3) | | | | | 1 (3.3) |
| 18 | | | | | | 1 (25.0) | | | | 1 (3.3) |
| Total SQP33 cumulative dose (mg/m$^2$) | | | | | | | | | | |
| Mean | 238.5 | 161.5 | 553.9 | 405.4 | 3692.1 | 4283 | 4157.5 | 2277.9 | 4206.1 | 2939.7 |
| SD | | | | 151.7 | 1542.71 | 4616.35 | 2633.99 | 1275.47 | 799.06 | 2421.29 |
| Median | 238.5 | 161.5 | 553.9 | 347.8 | 4226 | 2441.9 | 3710 | 2250.9 | 4686.8 | 2381.2 |
| (Min, Max) | (239, 239) | (162, 162) | (554, 554) | (291, 578) | (1953, 4897) | (1249, 11000) | (1905, 7305) | (499, 4248) | (3105, 4782) | (162, 11000) |
| Maximum SQP33 Dose Administered | | | | | | | | | | |
| 8 | 1 (100.0) | | | | | | | | | 1 (3.3) |
| 16 | | 1 (100.0) | | | | | | | | 1 (3.3) |
| 31 | | | 1 (100.0) | | | | | | | 1 (3.3) |
| 58 | | | | 3 (100.0) | | | | | | 3 (10.0) |

| | 10 mL SQL70 Biopolymer + SQP33 Dox Eq Dose (mg/m$^2$) | | | | | | | | | |
|---|---|---|---|---|---|---|---|---|---|---|
| | **0.38x (N=1)** | **0.76x (N=1)** | **1.53x (N=1)** | **2.8x (N=3)** | **4x (N=3)** | **6x (N=4)** | **8.8x (N=4)** | **12x (N=7)** | **15x (N=6)** | **Total (N=30)** |
| 84 | | | | | | | | | | |
| 85 | | | | | 1 (33.3) | | | | | 1 (3.3) |
| 125 | | | | | 1 (33.3) | 4 (100.0) | | | | 5 (16.7) |
| 185 | | | | | | | 2 (50.0) | | | 2 (6.7) |
| 186 | | | | | 1 (33.3) | | | | | 1 (3.3) |
| 192 | | | | | | | 2 (50.0) | | | 2 (6.7) |
| 249 | | | | | | | | 2 (28.6) | | 2 (6.7) |
| 250 | | | | | | | | 4 (57.1) | | 4 (13.3) |
| 253 | | | | | | | | 1 (14.3) | | 1 (3.3) |
| 311 | | | | | | | | | 1 (16.7) | 1 (3.3) |
| 312 | | | | | | | | | 1 (16.7) | 1 (3.3) |
| 315 | | | | | | | | | 1 (16.7) | 1 (3.3) |
| 322 | | | | | | | | | 2 (33.3) | 2 (6.7) |
| 326 | | | | | | | | | 1 (16.7) | 1 (3.3) |

Data Cutoff: 15 December 2022.
N = number of patients who received ≥1 dose of SQL70 biopolymer and 1 dose of SQP33 protodrug.
Dox Eq, doxorubicin molar equivalents; Max, maximum; Min, minimum; SD, standard deviation.

**Supplementary Table S4: SQ3370 administration and exposure for the Phase 1 Dose Escalation Group 2**

| | 20 mL SQL70 Biopolymer + SQP33 Dox Eq Dose (mg/m$^2$) | | |
|---|---|---|---|
| | **4x (N=5)** | **6x (N=3)** | **Total (N=8)** |
| Duration of SPQ3370 therapy (days) | | | |
| Mean | 42.6 | 49.3 | 45.1 |
| SD | 17.52 | 59.53 | 34.64 |
| Median | 46 | 26 | 36 |
| (Min, Max) | (26, 68) | (5, 117) | (5, 117) |
| Number of SQL70 cycles started | | | |
| 1 | -- | 1 (33.3%) | 1 (12.5%) |
| 2 | 2 (40.0%) | 1 (33.3%) | 3 (37.5%) |
| 3 | 2 (40.0%) | -- | 2 (25.0%) |
| 4 | 1 (20.0%) | -- | 1 (12.5%) |
| 5 | -- | -- | -- |
| 6 | | 1 (33.3%) | 1 (12.5%) |
| Average duration of SQP33 infusion (minutes) | | | |
| Mean | 30.8 | 30 | 30.5 |
| SD | 1.3 | 0 | 1.07 |
| Median | 30 | 30 | 30 |
| (Min, Max) | (30, 33) | (30, 30) | (30, 33) |
| Total SQP33 cumulative dose (mg/m$^2$) | | | |
| Mean | 1067.6 | 1866.8 | 1367.3 |
| SD | 401.32 | 1639.52 | 1367.3 |
| Median | 934.8 | 1249.0 | 1062.7 |
| (Min, Max) | 660.0, 1700.0 | 626.0, 3726.0 | 626.0, 3726.0 |
| Maximum SQP33 Dose Administered | | | |
| 8 | – | – | – |
| 16 | – | – | – |
| 31 | – | – | – |
| 58 | – | – | – |
| 84 | 1 (20.0%) | 1 (12.5%) | 1 (20.0%) |
| 85 | 4 (80.0%) | – | 4 (50.0%) |
| 125 | – | 3 (100.0%) | 3 (37.5%) |

Data Cutoff: 15 December 2022.
N = number of patients who received ≥1 dose of SQL70 biopolymer and 1 dose of SQP33 protodrug.
Dox Eq, doxorubicin molar equivalents; MedDRA, Medical Dictionary for Regulatory Activities; max, maximum; min, minimum; SD, standard deviation

**Supplementary Table S5: Pharmacokinetic parameters for Phase 1 (12x) and Phase 2a**

| Treatment arm | Day | SQL70 dose (mL) | SQP33 dose scheduled (mg/m$^2$) | Analyte | Variable | Units | N | Mean | SD | CV% | Median | Min | Max |
|---|---|---|---|---|---|---|---|---|---|---|---|---|---|
| Phase 1 | | | | | | | | | | | | | |
| Phase 1, 12x | 1 | 10 | 250 | Dox | $C_{max}$ | ng/mL | 7 | 1030 | 723.3 | 70.5 | 812 | 182 | 2180 |
| | | | | | $T_{max}$ | h | 7 | - | - | - | 0.583 | 0.567 | 4.5 |
| | | | | | $AUC_t$ | h*ng/mL | 7 | 2060 | 1655 | 80.3 | 1340 | 671 | 5100 |
| | | | | | $AUC_{tau}$ | h*ng/mL | 6 | 3440 | 2020 | 58.8 | 3550 | 1040 | 6650 |
| Phase 1, 12x | 1 | 10 | 250 | SQP 33 | $C_{max}$ | ng/mL | 7 | 24600 | 13980 | 56.7 | 28200 | 5.22 | 39900 |
| | | | | | $T_{max}$ | h | 7 | - | - | - | 0.583 | 0.567 | 1.08 |
| | | | | | $AUC_t$ | h*ng/mL | 7 | 19100 | 12040 | 62.9 | 23200 | 4.62 | 30700 |
| | | | | | $AUC_{tau}$ | h*ng/mL | 6 | 22500 | 9621 | 42.7 | 25500 | 6910 | 31500 |
| Phase 1, 12x | 5 | 10 | 250 | Dox | $C_{max}$ | ng/mL | 6 | 60.4 | 20.65 | 34.2 | 64.6 | 32.7 | 83.2 |
| | | | | | $T_{max}$ | h | 6 | - | - | - | 0.583 | 0.567 | 0.667 |
| | | | | | $AUC_t$ | h*ng/mL | 6 | 117 | 41.33 | 35.3 | 122 | 68.4 | 173 |
| | | | | | $AUC_{tau}$ | h*ng/mL | 6 | 233 | 108.3 | 46.4 | 255 | 100 | 353 |
| Phase 1, 12x | 5 | 10 | 250 | SQP 33 | $C_{max}$ | ng/mL | 6 | 31800 | 12960 | 40.8 | 35000 | 8670 | 46400 |
| | | | | | $T_{max}$ | h | 6 | - | - | - | 0.583 | 0.567 | 0.667 |
| | | | | | $AUC_t$ | h*ng/mL | 6 | 30400 | 17640 | 58 | 26800 | 7130 | 57900 |

| Treatment arm | Day | SQL70 dose (mL) | SQP33 dose scheduled (mg/m$^2$) | Analyte | Variable | Units | N | Mean | SD | CV% | Median | Min | Max |
|---|---|---|---|---|---|---|---|---|---|---|---|---|---|
| | | | | | $AUC_{tau}$ | h*ng/mL | 6 | 32500 | 20810 | 64.1 | 27300 | 7200 | 67400 |
| Phase 2a | | | | | | | | | | | | | |
| Dose Expansion A (Low Dose) | 1 | 10 | 185 | Dox | $C_{max}$ | ng/mL | 1 | 698 | - | - | - | - | - |
| | | | | | $T_{max}$ | h | 1 | - | - | - | 0.58 | - | - |
| | | | | | $AUC_{last}$ | h*ng/mL | 1 | 6541.53 | - | - | - | - | - |
| | | | | | $AUC_{inf}$ | h*ng/mL | 1 | 7003.08 | - | - | - | - | - |
| Dose Expansion A (Low Dose) | 1 | 10 | 185 | SQP 33 | $C_{max}$ | ng/mL | 1 | 29200 | - | - | - | - | - |
| | | | | | $T_{max}$ | h | 1 | - | - | - | 0.58 | - | - |
| | | | | | $AUC_{last}$ | h*ng/mL | 1 | 27695.67 | - | - | - | - | - |
| | | | | | $AUC_{inf}$ | h*ng/mL | 1 | 27707.05 | - | - | - | - | - |
| Dose Expansion A (Low Dose) | 5 | 10 | 185 | Dox | $C_{max}$ | ng/mL | 1 | 70.5 | - | - | - | - | - |
| | | | | | $T_{max}$ | h | 1 | - | - | - | 0.58 | - | - |
| | | | | | $AUC_{inf}$ | h*ng/mL | 1 | 376.11 | - | - | - | - | - |
| | | | | | $AUC_{last}$ | h*ng/mL | 1 | 179.16 | - | - | - | - | - |
| Dose Expansion A (Low Dose) | 5 | 10 | 185 | SQP 33 | $C_{max}$ | ng/mL | 1 | 26900 | - | - | - | - | - |
| | | | | | $T_{max}$ | h | 1 | 0.58 | - | - | 0.58 | - | - |

| Treatment arm | Day | SQL70 dose (mL) | SQP33 dose scheduled (mg/m$^2$) | Analyte | Variable | Units | N | Mean | SD | CV% | Median | Min | Max |
|---|---|---|---|---|---|---|---|---|---|---|---|---|---|
| | | | | | $AUC_{inf}$ | h*ng/mL | 1 | 33653.97 | - | - | - | - | - |
| | | | | | $AUC_{last}$ | h*ng/mL | 1 | 32464.26 | - | - | - | - | - |
| Expansion Group 1: Extremity STS - 5 Day | 1 | 20 | 250 | Dox | $C_{max}$ | ng/mL | 2 | 245.05 | 270.04 | 110.2 | 245.05 | 54.1 | 436 |
| | | | | | $T_{max}$ | h | 2 | 0.75 | 0 | 0 | 0.75 | 0.75 | 0.75 |
| | | | | | $AUC_{inf}$ | h*ng/mL | 2 | 2512.02 | 2773.47 | 110.41 | 2512.02 | 550.88 | 4473.16 |
| | | | | | $AUC_{last}$ | h*ng/mL | 2 | 2420.11 | 2677.51 | 110.64 | 2420.11 | 526.83 | 4313.4 |
| Expansion Group 1: Extremity STS - 5 Day | 1 | 20 | 250 | SQP 33 | $C_{max}$ | ng/mL | 2 | 13520 | 8032.73 | 59.41 | 13520 | 7840 | 19200 |
| | | | | | $T_{max}$ | h | 2 | - | - | - | 0.75 | 0.75 | 0.75 |
| | | | | | $AUC_{inf}$ | h*ng/mL | 0 | - | - | - | - | - | - |
| | | | | | $AUC_{last}$ | h*ng/mL | 2 | 19500.04 | 12346.74 | 63.32 | 19500.04 | 10769.58 | 28230.5 |
| Expansion Group 1: Extremity STS - 5 Day | 5 | 20 | 250 | Dox | $C_{max}$ | ng/mL | 2 | 42.05 | 25.95 | 61.71 | 42.05 | 23.7 | 60.4 |
| | | | | | $T_{max}$ | h | 2 | - | - | - | 0.81 | 0.78 | 0.83 |
| | | | | | $AUC_{inf}$ | h*ng/mL | 0 | - | - | - | - | - | - |
| | | | | | $AUC_{last}$ | h*ng/mL | 2 | 108.93 | 77.56 | 71.2 | 108.93 | 54.08 | 163.77 |
| Expansion Group 1: Extremity STS - 5 Day | 5 | 20 | 250 | SQP 33 | $C_{max}$ | ng/mL | 2 | 20450 | 14495.69 | 70.88 | 20450 | 10200 | 30700 |
| | | | | | $T_{max}$ | h | 2 | - | - | - | 0.81 | 0.78 | 0.83 |
| | | | | | $AUC_{inf}$ | h*ng/mL | 0 | - | - | - | - | - | - |

| Treatment arm | Day | SQL70 dose (mL) | SQP33 dose scheduled (mg/m²) | Analyte | Variable | Units | N | Mean | SD | CV% | Median | Min | Max |
|---|---|---|---|---|---|---|---|---|---|---|---|---|---|
| | | | | | $AUC_{last}$ | h*ng/mL | 2 | 29151.23 | 20860.39 | 71.56 | 29151.23 | 14400.7 | 43901.75 |
| Expansion Group 2A: Unresectable STS - 5 day | 1 | 10 | 250 | Dox | $C_{max}$ | ng/mL | 6 | 672.83 | 381.86 | 56.75 | 790.5 | 101 | 1070 |
| | | | | | $T_{max}$ | h | 6 | - | - | - | 0.75 | 0 | 0.85 |
| | | | | | $AUC_{inf}$ | h*ng/mL | 5 | 2659.08 | 1060.6 | 39.89 | 2783.81 | 932.5 | 3680.26 |
| | | | | | $AUC_{last}$ | h*ng/mL | 6 | 2368.5 | 1049.28 | 44.3 | 2593.87 | 896.87 | 3581.74 |
| Expansion Group 2A: Unresectable STS - 5 day | 1 | 10 | 250 | SQP 33 | $C_{max}$ | ng/mL | 6 | 18416.67 | 6372.26 | 34.6 | 17150 | 11900 | 28500 |
| | | | | | $T_{max}$ | h | 6 | - | - | - | 0.75 | 0.68 | 0.83 |
| | | | | | $AUC_{inf}$ | h*ng/mL | 0 | - | - | - | - | - | - |
| | | | | | $AUC_{last}$ | h*ng/mL | 6 | 42899.68 | 90521.8 | 211.01 | 6309.04 | 4462.5 | 227659.2 |
| Expansion Group 2A: Unresectable STS - 5 day | 5 | 10 | 250 | Dox | $C_{max}$ | ng/mL | 6 | 28.65 | 6.75 | 23.57 | 26.45 | 21 | 40 |
| | | | | | $T_{max}$ | h | 6 | - | - | - | 0.79 | 0.75 | 0.87 |
| | | | | | $AUC_{inf}$ | h*ng/mL | 0 | - | - | - | - | - | - |
| | | | | | $AUC_{last}$ | h*ng/mL | 6 | 77.27 | 18.82 | 24.36 | 76.09 | 59.51 | 111.78 |
| Expansion Group 2A: Unresectable STS - 5 day | 5 | 10 | 250 | SQP 33 | $C_{max}$ | ng/mL | 6 | 15101.67 | 6428.53 | 42.57 | 14550 | 6610 | 24600 |
| | | | | | $T_{max}$ | h | 6 | - | - | - | 0.8 | 0.75 | 2.08 |
| | | | | | $AUC_{inf}$ | h*ng/mL | 0 | - | - | - | - | - | - |
| | | | | | $AUC_{last}$ | h*ng/mL | 6 | 22725.57 | 8372.14 | 36.84 | 23798.61 | 10710.3 | 32657.1 |

| Treatment arm | Day | SQL70 dose (mL) | SQP33 dose scheduled (mg/m$^2$) | Analyte | Variable | Units | N | Mean | SD | CV% | Median | Min | Max |
|---|---|---|---|---|---|---|---|---|---|---|---|---|---|
| Expansion Group 2B: Unresectable STS - 3 day | 1 | 10 | 250 | Dox | $C_{max}$ | ng/mL | 6 | 780.03 | 732.47 | 93.9 | 688 | 75.2 | 1880 |
| | | | | | $T_{max}$ | h | 6 | - | - | - | 0.81 | 0.77 | 0.92 |
| | | | | | $AUC_{inf}$ | h*ng/mL | 4 | 5283.58 | 2815.16 | 53.28 | 5248.86 | 1881.47 | 8755.14 |
| | | | | | $AUC_{last}$ | h*ng/mL | 6 | 3886.78 | 2971.11 | 76.44 | 3646.85 | 256.74 | 8552.82 |
| Expansion Group 2B: Unresectable STS - 3 day | 1 | 10 | 250 | SQP 33 | $C_{max}$ | ng/mL | 6 | 17198.3 3 | 5880.55 | 34.19 | 17100 | 7990 | 25600 |
| | | | | | $T_{max}$ | h | 6 | - | - | - | 0.81 | 0.77 | 0.92 |
| | | | | | $AUC_{inf}$ | h*ng/mL | 0 | - | - | - | - | - | - |
| | | | | | $AUC_{last}$ | h*ng/mL | 6 | 19934.6 4 | 10818.24 | 54.27 | 20587.9 3 | 6976.67 | 35361.64 |
| Expansion Group 2B: Unresectable STS - 3 day | 3 | 10 | 500 | Dox | $C_{max}$ | ng/mL | 6 | 54.4 | 22.59 | 41.52 | 45.65 | 36.6 | 96.9 |
| | | | | | $T_{max}$ | h | 6 | 1.24 | 0.41 | 32.97 | 1.28 | 0.75 | 1.92 |
| | | | | | $AUC_{inf}$ | h*ng/mL | 0 | | - | - | - | - | - |
| | | | | | $AUC_{last}$ | h*ng/mL | 6 | 163.33 | 43.21 | 26.46 | 155.81 | 99.44 | 218.24 |
| Expansion Group 2B: Unresectable STS - 3 day | 3 | 10 | 500 | SQP 33 | $C_{max}$ | ng/mL | 6 | 44350 | 10301.02 | 23.23 | 44400 | 27200 | 57300 |
| | | | | | $T_{max}$ | h | 6 | - | - | - | 1.28 | 0.75 | 1.92 |
| | | | | | $AUC_{inf}$ | h*ng/mL | 0 | - | - | - | - | - | - |
| | | | | | $AUC_{last}$ | h*ng/mL | 6 | 87270.6 9 | 33515.8 | 38.4 | 76743.1 9 | 44752.33 | 131671 |

Abbreviations: $AUC_{last}$ = Area Under Curve from time zero through the last quantifiable sample; $AUC_{inf}$ = Area Under Curve from time zero extrapolated to infinity; CV = coefficient of variance; $C_{max}$ = Observed maximum plasma concentration; min = minimal; max = maximal; SD = standard deviation; $T_{1/2}$ = Terminal elimination half-life; $T_{max}$ = Time to maximum concentration.

### Supplementary Table S6: TEAEs in the Phase 1 Dose Escalation Group 1 cohort (Safety Population)

| | Dose Escalation: 10 mL SQL70 Biopolymer + SQP33 Dox Eq Dose (mg/m2) | | | | | | | | | |
|---|---|---|---|---|---|---|---|---|---|---|
| | **0.38x (N=1)** | **0.76x (N=1)** | **1.53x (N=1)** | **2.8x (N=3)** | **4x (N=3)** | **6x (N=4)** | **8.8x (N=4)** | **12x (N=7)** | **15x (N=6)** | **Total (N=30)** |
| Number of TEAEs reported | 8 | 3 | 15 | 31 | 31 | 22 | 47 | 80 | 53 | 290 |
| Patients with TEAEs, n (%) | | | | | | | | | | |
| Any TEAEs | 1 (100.0% | 1 (100.0) | 1 (100.0) | 3 (100.0) | 3 (100.0) | 4 (100.0) | 4 (100.0) | 7 (100.0) | 6 (100.0) | 30 (100.0) |
| TEAEs occurring in ≥ 2 patients | | | | | | | | | | |
| Nausea | 0 (0.0%) | 1 (100.0%) | 1 (100.0%) | 2 (66.7%) | 2 (66.7%) | 3 (75.0%) | 1 (25.0%) | 4 (57.1%) | 3 (50.0%) | 17 (56.7%) |
| Constipation | 1 (100.0%) | 0 (0.0%) | 1 (100.0%) | 1 (33.3%) | 0 (0.0%) | 1 (25.0%) | 1 (25.0%) | 2 (28.6%) | 2 (33.3%) | 9 (30.0%) |
| Stomatitis | 0 (0.0%) | 0 (0.0%) | 0 (0.0%) | 0 (0.0%) | 0 (0.0%) | 0 (0.0%) | 2 (50.0%) | 2 (28.6%) | 1 (16.7%) | 5 (16.7%) |
| Fatigue | 1 (100.0%) | 0 (0.0%) | 1 (100.0%) | 3 (100.0%) | 1 (33.3%) | 1 (25.0%) | 1 (25.0%) | 3 (42.9%) | 4 (66.7%) | 15 (50.0%) |
| Pyrexia | 0 (0.0%) | 0 (0.0%) | 0 (0.0%) | 2 (66.7%) | 1 (33.3%) | 0 (0.0%) | 0 (0.0%) | 2 (28.6%) | 0 (0.0%) | 5 (16.7%) |
| Decreased Appetite | 0 (0.0%) | 0 (0.0%) | 0 (0.0%) | 0 (0.0%) | 0 (0.0%) | 0 (0.0%) | 1 (25.0%) | 3 (42.9%) | 3 (50.0%) | 7 (23.3%) |
| Covid-19 | 0 (0.0%) | 0 (0.0%) | 0 (0.0%) | 0 (0.0%) | 0 (0.0%) | 0 (0.0%) | 2 (50.0%) | 3 (42.9%) | 0 (0.0%) | 5 (16.7%) |
| Anaemia | 0 (0.0%) | 0 (0.0%) | 0 (0.0%) | 1 (33.3%) | 1 (33.3%) | 1 (25.0%) | 2 (50.0%) | 1 (14.3%) | 1 (16.7%) | 7 (23.3%) |
| Dyspnoea | 0 (0.0%) | 0 (0.0%) | 0 (0.0%) | 1 (33.3%) | 1 (33.3%) | 1 (25.0%) | 0 (0.0%) | 1 (14.3%) | 1 (16.7%) | 5 (16.7%) |
| Alopecia | 0 (0.0%) | 0 (0.0%) | 0 (0.0%) | 0 (0.0%) | 0 (0.0%) | 0 (0.0%) | 1 (25.0%) | 1 (14.3%) | 3 (50.0%) | 5 (16.7%) |
| TEAEs related to study drug | 0 (0) | 1 (100.0) | 1 (100.0) | 3 (100.0) | 3 (100.0) | 3 (75.0) | 3 (75.0) | 5 (71.4) | 5 (83.3) | 24 (80.0) |
| Serious TEAE | 1 (100.0) | 0 (0) | 1 (100.0) | 2 (66.7) | 2 (66.7) | 0 (0) | 2 (50.0) | 2 (28.6) | 0 (0) | 10 (33.3) |
| TEAE leading to study discontinuation | 0 (0) | (0 (0) | 0 (0) | 0 (0) | 0 (0) | 1 (25.0) | 0 (0) | 2 (28.6) | 0 (0) | 3 (10.0) |
| Cardiac failure | 0 (0) | 0 (0) | 0 (0) | 0 (0) | 0 (0) | 0 (0) | 0 (0) | 1 (14.3) | 0 (0) | 1 (3.3) |
| Fatigue | 0 (0) | 0 (0) | 0 (0) | 0 (0) | 0 (0) | 0 (0) | 0 (0) | 1 (14.3) | 0 (0) | 1 (3.3) |
| Dyspnoe | 0 (0) | 0 (0) | 0 (0) | 0 (0) | 0 (0) | 1 (25.0) | 0 (0) | 0 (0) | 0 (0) | 1 (3.3) |
| DLTs | 0 (0) | 0 (0) | 0 (0) | 0 (0) | 0 (0) | 0 (0) | 0 (0) | 0 (0) | 0 (0) | 0 (0) |
| Grade 3 or 4 TEAEs | 1 (100.0) | 0 (0) | 1 (100.0) | 1 (33.3) | 1 (33.3) | 1 (25.0) | 2 (50.0) | 4 (57.1) | 1 (16.7) | 12 (40.0) |
| Death | 1 (100.0) | 0 (0) | 0 (0) | 0 (0) | 0 (0) | 0 (0) | 0 (0) | 0 (0) | 0 (0) | 1 (3.3) |
| Patients with AESIs, n (%) | 0 (0) | 0 (0) | 0 (0) | 0 (0) | 0 (0) | 0 (0) | 2 (50.0) | 3 (42.9) | 0 (0) | 5 (16.7) |
| Covid infections | 0 (0) | 0 (0) | 0 (0) | 0 (0) | 0 (0) | 0 (0) | 2 (50.0) | 3 (42.9) | 0 (0) | 5 (16.7) |
| Cardiac failure | 0 (0) | 0 (0) | 0 (0) | 0 (0) | 0 (0) | 0 (0) | 0 (0) | 1 (14.3) | 0 (0) | 1 (3.3) |

| | Dose Escalation: 10 mL SQL70 Biopolymer + SQP33 Dox Eq Dose (mg/m2) | | | | | | | | | |
|---|---|---|---|---|---|---|---|---|---|---|
| | **0.38x (N=1)** | **0.76x (N=1)** | **1.53x (N=1)** | **2.8x (N=3)** | **4x (N=3)** | **6x (N=4)** | **8.8x (N=4)** | **12x (N=7)** | **15x (N=6)** | **Total (N=30)** |
| Patients with adverse events of myelosuppression severity, n (%) | 0 (0) | 0 (0) | 0 (0) | 0 (0) | 0 (0) | 0 (0) | 2 (50.0) | 3 (42.9) | 0 (0) | 5 (16.7) |
| Anemia | | | | | | | | | | |
| Grade 1 | 0 (0) | 0 (0) | 0 (0) | 0 (0) | 0 (0) | 0 (0) | 1 (25.0) | 0 (0) | 0 (0) | 1 (3.3) |
| Grade 2 | 0 (0) | 0 (0) | 0 (0) | 1 (33.3) | 0 (0) | 0 (0) | 0 (0) | 0 (0) | 0(0) | 1 (3.3) |
| Grade 3 | 0 (0) | 0 (0) | 0 (0) | 0 (0) | 1 (33.3) | 1 (25.0) | 1 (25.0) | 0 (0) | 1 (16.7) | 4 (13.3) |
| Grade 4 | 0 (0) | 0 (0) | 0 (0) | 0 (0) | 0 (0) | 0 (0) | 0 (0) | 1 (14.3) | 0 (0) | 1 (3.3) |
| Grade 5 | 0 (0) | 0 (0) | 0 (0) | 0 (0) | 0 (0) | 0 (0) | 0 (0) | 0 (0) | 0 (0) | 0 (0) |
| Neutropenia | | | | | | | | | | |
| Grade 1 | 0 (0) | 0 (0) | 0 (0) | 0 (0) | 0 (0) | 1 (25.0) | 0 (0) | 0 (0) | 2 (33.3) | 3 (10.0) |
| Grade 2 | 0 (0) | 0 (0) | 0 (0) | 1 (33.3) | 1 (33.3) | 0 (0) | 0 (0) | 1 (14.3) | 0 (0) | 2 (10.0) |
| Grade 3 | 0 (0) | 0 (0) | 0 (0) | 0 (0) | 0 (0) | 0 (0) | 0 (0) | 1 (14.3) | 0 (0) | 1 (3.3) |
| Grade 4 | 0 (0) | 0 (0) | 0 (0) | 0 (0) | 0 (0) | 0 (0) | 1 (25.0) | 1 (14.3) | 0 (0) | 2 (6.7) |
| Grade 5 | 0 (0) | 0 (0) | 0 (0) | 0 (0) | 0 (0) | 0 (0) | 0 (0) | 0 (0) | 0 (0) | 0 (0) |
| Thrombocytopenia | | | | | | | | | | |
| Grade 1 | 0 (0) | 1 (100.0) | 0 (0) | 1 (33.3) | 1 (33.3) | 0 (0) | 0 (0) | 1 (14.3) | 1 (16.7) | 5 (16.7) |
| Grade 2 | 0 (0) | 0 (0) | 0 (0) | 0 (0) | 0 (0) | 0 (0) | 0 (0) | 0 (0) | 0 (0) | 0 (0) |
| Grade 3 | 0 (0) | 0 (0) | 0 (0) | 0 (0) | 0 (0) | 0 (0) | 0 (0) | 0 (0) | 0 (0) | 0 (0) |
| Grade 4 | 0 (0) | 0 (0) | 0 (0) | 0 (0) | 0 (0) | 0 (0) | 1 (25.0) | 1 (14.3) | 0 (0) | 2 (6.7) |
| Grade 5 | 0 (0) | 0 (0) | 0 (0) | 0 (0) | 0 (0) | 0 (0) | 0 (0) | 0 (0) | 0 (0) | 0 (0) |

Data Cutoff: 15 December 2022.
N = number of patients who received ≥1 dose of SQL70 biopolymer and 1 dose of SQP33 protodrug.
TEAEs were coded using MedDRA Version 24.1.
DLT, dose-limiting toxicity; Dox Eq, doxorubicin molar equivalents; MedDRA, Medical Dictionary for Regulatory Activities; max, maximum; min, minimum; SD, standard deviation; TEAE, treatment emergent adverse event.

**Supplementary Table S7: TEAEs in the Phase 1 Dose Escalation Group 2 cohort (safety population)**

| | 20 mL SQL70 Biopolymer + SQP33 Dox Eq Dose (mg/m$^2$) | | |
|---|---|---|---|
| | **4x (N = 5)** | **6x (N = 3)** | **Total (N = 8)** |
| Number of TEAEs reported | 44 | 19 | 63 |
| Patients with TEAEs, n (%) | | | |
| Any TEAEs | 5 (100.0) | 3 (100.0) | 8 (100.0) |
| TEAEs occurring in ≥ 2 patients | | | |
| Nausea | 2 (40.0) | 2 (66.7) | 4 (50.0) |
| Diarrhea | 2 (40.0) | 0 (0) | 2 (25.0) |
| Fatigue | 1 (20.0) | 1 (33.3) | 2 (25.0) |
| Decreased appetite | 2 (40.0) | 2 (66.7) | 4 (50.0) |
| Anemia | 4 (80.0) | 0 (0) | 4 (50.0) |
| Neutropenia | 2 (40.0) | 0 (0) | 2 (25.0) |
| Dizziness | 1 (20.0) | 1 (33.3) | 2 (25.) |
| Related TEAEs | 4 (80.0) | 2 (66.7) | 6 (75.0) |
| Serious TEAE | 1 (20.0) | 1 (33.3) | 2 (25.0) |
| TEAE leading to study discontinuation | 0 (0) | 0 (0) | 0 (0) |
| Cardiac failure | 0 (0) | 0 (0) | 0 (0) |
| Fatigue | 0 (0) | 0 (0) | 0 (0) |
| Dyspnoe | 0 (0) | 0 (0) | 0 (0) |
| DLTs | 0 (0) | 0 (0) | 0 (0) |
| Grade 3 or 4 TEAEs | 3 (60.0) | 1 (33.3) | 4 (50.0) |
| Death | 0 (0) | 0 (0) | 0 (0) |
| Patients with AESIs, n (%) | 1 (20.0) | 0 (0) | 1 (12.5) |
| Covid infections | 1 (2.0) | 0 (0) | 1 (12.5) |
| Cardiac failure | 0 (0) | 0 (0) | 0 (0) |
| Patients with adverse events of myelosuppression by severity, n (%) | 1 (20.0) | 0 (0) | 1 (12.5) |
| Anemia | | | |
| Grade 1 | 0 (0) | 0 (0) | 0 (0) |
| Grade 2 | 1 (20.0) | 0 (0) | 1 (12.5) |
| Grade 3 | 3 (60.0) | 0 (0) | 3 (37.5) |
| Grade 4 | 0 (0) | 0 (0) | 0 (0) |
| Grade 5 | 0 (0) | 0 (0) | 0 (0) |
| Neutropenia | | | |
| Grade 1 | 0 (0) | 0 (0) | 0 (0) |
| Grade 2 | 0 (0) | 0 (0) | 0 (0) |
| Grade 3 | 1 (20.0) | 0 (0) | 1 (12.5) |
| Grade 4 | 1 (20.0) | 0 (0) | 1 (12.5) |
| Grade 5 | 0 (0) | 0 (0) | 0 (0) |
| Thrombocytopenia | | | |
| Grade 1 | 0 (0) | 0 (0) | 0 (0) |
| Grade 2 | 0 (0) | 0 (0) | 0 (0) |
| Grade 3 | 1 (20.0) | 0 (0) | 1 (12.5) |
| Grade 4 | 0 (0) | 0 (0) | 0 (0) |
| Grade 5 | 0 (0) | 0 (0) | 0 (0) |

Data Cutoff: 15 December 2022.
N = number of patients who received ≥1 dose of SQL70 biopolymer and 1 dose of SQP33 protodrug.
TEAEs were coded using MedDRA Version 24.1.

DLT, dose-limiting toxicity; Dox Eq, doxorubicin molar equivalents; MedDRA, Medical Dictionary for Regulatory Activities; max, maximum; min, minimum; SD, standard deviation; TEAE, treatment emergent adverse event.
